# Diagnosis of Multiple Sclerosis Using Multimodal Deep Learning Integrating Lesion and Normal-Appearing White Matter: A Retrospective Study with International Multicentre External Validation

**DOI:** 10.64898/2026.03.04.26347460

**Authors:** Jiajian Ma, Valentin Stepanov, Wushuang Rui, Hsuan-Chih Chen, Maciej Lis, Aleksandra Stanek, Tomasz Puto, Michael Lan, Jenny Chen, Timothy Liu, Roshni Patel, Matthew Breen, Matthew Lee, Katharina Eikermann-Haerter, Timothy M. Shepherd, Dmitry S. Novikov, Kimberly A. O’Neill, Els Fieremans, Yiqiu Shen

## Abstract

Current multiple sclerosis (MS) diagnosis relies primarily on focal white matter lesions (WMLs), which are frequently mimicked by other conditions. Normal-appearing white matter (NAWM) harbours complementary pathological information but remains clinically underutilised because NAWM alterations are macroscopically occult on routine scans and require non-routine quantitative imaging to visualise. Here, we show that NAWM-related diagnostic information can be recovered from routine structural MRI using a cross-modal deep-learning model. We developed DeepMS, a model co-trained on diffusion and structural MRI that operates solely on structural MRI at deployment. DeepMS achieved ROC-AUCs of 0.968 internally (n=837) and 0.940–0.974 across two international external cohorts (n=293 and n=1,756). In a multi-reader study, DeepMS outperformed the 2024 McDonald criteria imaging biomarkers. DeepMS retained robust performance after digital lesion removal and exhibited NAWM-dominant activation maps. Combined with established imaging biomarkers, DeepMS improved sensitivity (92.1% vs 74.8%) while maintaining high specificity (95.6% vs 92.3%) compared with corresponding biomarker composite based on the 2024 McDonald criteria. By decoding latent NAWM signals from routine scans and integrating them with WML features, this framework can potentially advance MS diagnosis beyond the current lesion-centric paradigm.

## Introduction

Multiple sclerosis (MS) is a leading cause of non-traumatic neurological disability in young adults^1^, and accurate diagnosis is critical for the timely initiation of disease-modifying therapy and appropriate clinical management^2^. Current MS diagnostic criteria rely heavily on MRI biomarkers derived from focal white matter lesions (WMLs), such as the dissemination in space and time (DIS/DIT)^3^. However, these lesion patterns are not specific to MS and frequently appear in other neurological disorders, such as cerebral small vessel disease, migraine, and other inflammatory demyelinating disorders^4^. Although the 2024 revision of the McDonald criteria further incorporated more pathophysiologically specific imaging biomarkers^3^, including the central vein sign (CVS)^5^ and paramagnetic rim lesions (PRLs)^6^, these markers still exhibit limited sensitivity on routine clinical imaging, and their absence cannot reliably exclude MS^3^.

Consequently, MS misdiagnosis remains a persistent clinical problem under the current lesion-centred diagnostic framework: approximately 10–20% of patients referred for MS diagnosis are ultimately found to have alternative conditions^7–9^, leading to inappropriate immunosuppressive treatment and unnecessary patient anxiety^4^.

Beyond focal WMLs, diffuse microstructural injury within the normal-appearing white matter (NAWM) represents another pathological hallmark of MS^10^. Previous studies have shown that NAWM can harbour clinically relevant pathology, including axonal degeneration and microglial activation^10^, and that NAWM abnormalities are associated with disease activity^11^, functional impairment^12^, and prognosis^13^. Growing evidence further indicates that quantitative measures derived from the NAWM may aid in differentiating MS from radiological mimics^14–16^. However, NAWM-related diagnostic information has not been translated into clinical practice for MS diagnosis, owing to two major barriers. First, an MS-specific NAWM signature has not yet been established^3,17^. Second, detection of NAWM abnormalities typically requires quantitative analysis derived from specialised acquisitions, such as multi-shell diffusion MRI (dMRI)^12,18^, which are not part of routine clinical workflows^3,17^. A critical clinical challenge remains: decoding NAWM-derived diagnostic signals from routine MRI to complement established lesion-centric biomarkers.

Multi-shell dMRI can sensitively capture microstructural injury within the NAWM in MS by detecting alterations in microscopic water displacement, thereby providing critical clues to NAWM-related diagnostic information^19–21^. However, these specialised scans are rarely available at the point of care^3,17^. Multimodal deep learning offers a potential route to incorporate this advanced signal into standard practice. We hypothesized that by training a deep neural network on paired dMRI and routine structural MRI (sMRI), the model would learn to associate the microstructural damage visible on dMRI with subtle correlates hidden within the standard sMRI.

In this study, we developed DeepMS, a deep-learning model for MS diagnosis designed to extract both lesional and latent NAWM pathology from routine sMRI. Through paired dMRI– sMRI training, DeepMS learns to detect NAWM-related pathology from routine sMRI alone at deployment, converting underused NAWM information into clinically usable diagnostic signals (Fig. 1a). Across diverse, multicentre international cohorts, we demonstrate that DeepMS captures diagnostically relevant NAWM-related information from routine sMRI, exhibits robust generalisability, and provides complementary diagnostic value that substantially improves upon the established imaging biomarkers of the 2024 McDonald criteria^3^.

**Fig. 1:**
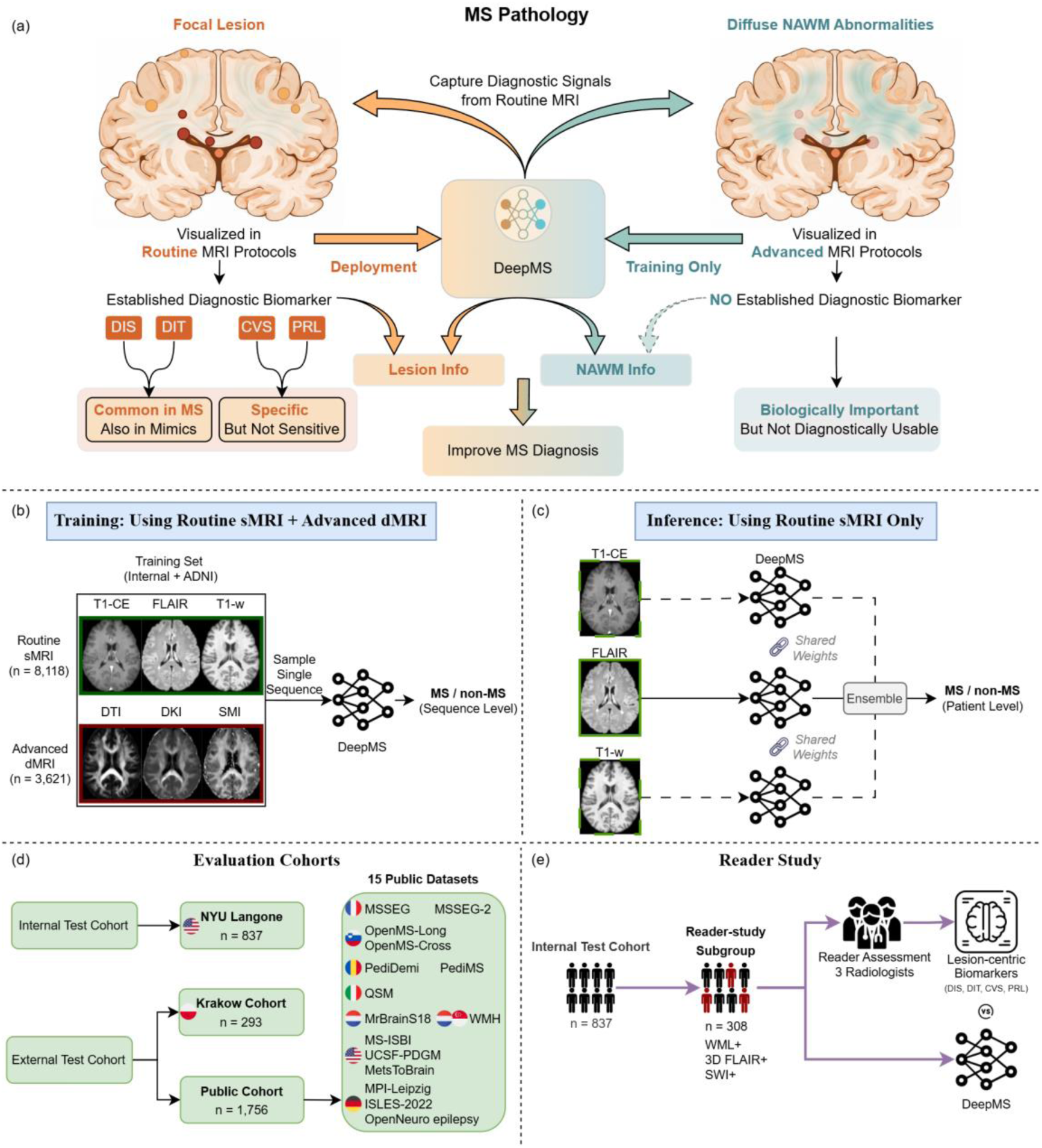
Study design of the DeepMS model. **a,** DeepMS was designed to capture diagnostic signals from both focal MS lesions visible on routine MRI and diffuse abnormalities in normal-appearing white matter detectable by advanced MRI, while enabling deployment using routine MRI alone. **b,** During training, DeepMS used routine structural MRI sequences, including T1-CE, FLAIR, and T1-w, together with advanced diffusion MRI sequences, including DTI, DKI, and SMI, to predict MS versus non-MS. **c,** During inference, only routine structural MRI sequences were used, with sequence-level predictions ensembled to generate a patient-level diagnosis. **d,** Model performance was evaluated in an internal test cohort and independent external cohorts, including the Krakow cohort and 15 public datasets. **e,** A reader study compared DeepMS with radiologist assessment based on lesion-centric imaging biomarkers, including DIS, DIT, CVS, and PRL. CVS, central vein sign; DIS, dissemination in space; DIT, dissemination in time; DKI, diffusional kurtosis imaging; dMRI, diffusion MRI; DTI, diffusion tensor imaging; NAWM, normal-appearing white matter; PRL, paramagnetic rim lesion; SMI, standard model imaging;

## Results

### Study overview and cohorts

DeepMS was initialized with pre-trained weights from a Swin-UNETR based foundation model^22,23^ and fine-tuned on retrospectively collected multimodal MRI data from NYU Langone Health and ADNI^24^, comprising 8,118 sMRI scans and 3,621 multi-shell dMRI scans from 7,382 participants (Fig. 1b). At inference, DeepMS generated a patient-level diagnostic score from fluid-attenuated inversion recovery (FLAIR) alone or in combination with other routine structural MRI sequences, including T1-weighted (T1-w) and contrast-enhanced T1-weighted (T1-CE) imaging, serving as a biomarker for MS diagnosis (Fig. 1c).

We evaluated DeepMS in an internal NYU Langone Health test cohort (n=837; 138 MS, 699 non-MS; 326 non-MS with WMLs), an external cohort from the 5th Military Hospital with Polyclinic, Krakow, Poland (n=293; 63 MS, 230 non-MS; 113 non-MS with WMLs), and a public multi-dataset external cohort spanning the USA25–27, Germany^28–30^, France31,32, Italy33, Netherlands34,35, Slovenia36,37, Romania38,39, and Singapore^35^ (n=1,756; 263 MS, 1,493 non-MS; 461 non-MS with WMLs) (Fig. 1d). Across cohorts, non-MS cases included diverse lesion-positive MS mimics, providing a clinically representative and diagnostically challenging evaluation setting. Detailed demographic and imaging characteristics are summarised in Table 1, Extended Data Fig. 1, and Extended Data Tables 1 and 2.

**Table 1.**
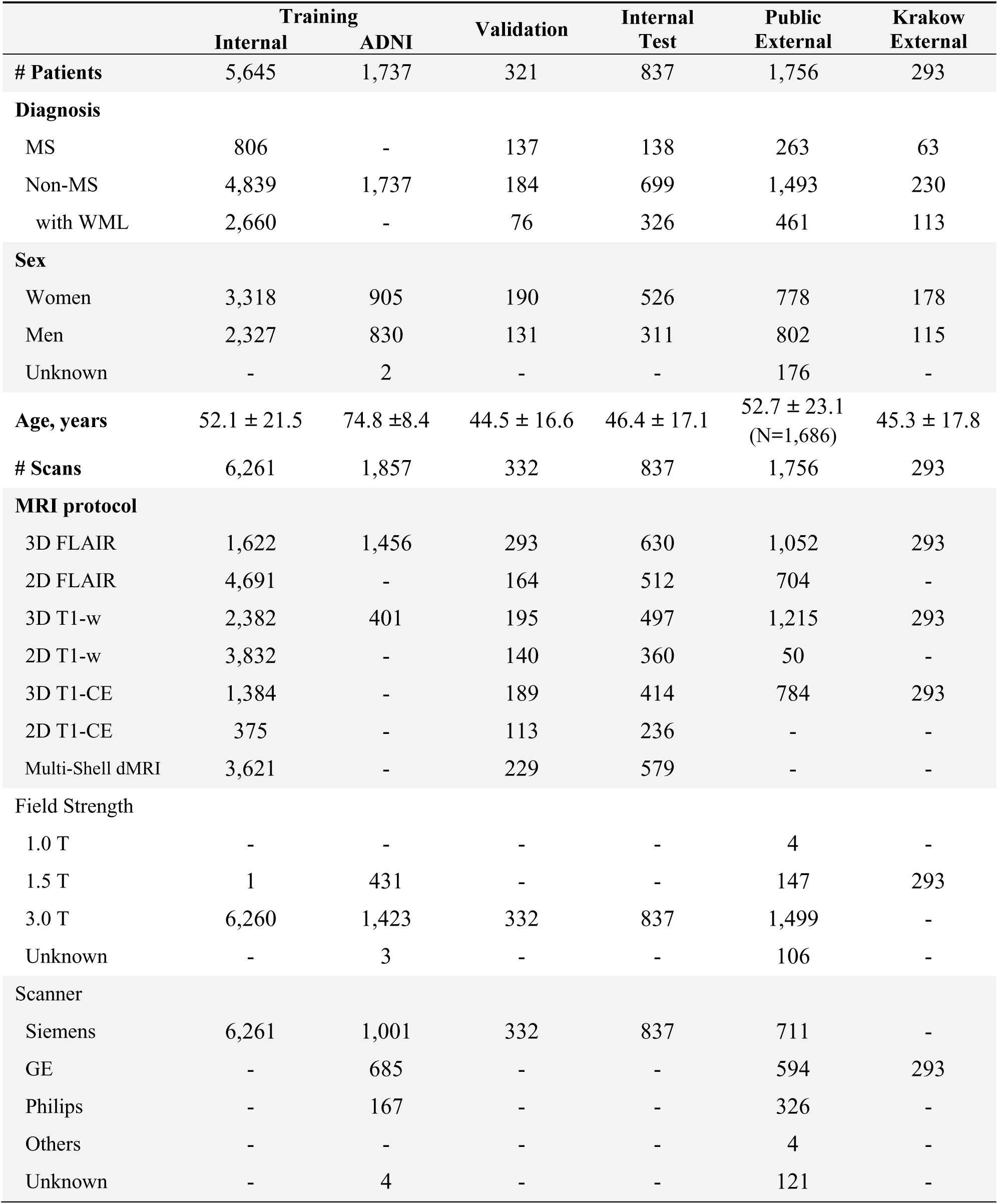
Cohort characteristics.

|  | Training |  | Validation | Internal Test | Public External | Krakow External |
| --- | --- | --- | --- | --- | --- | --- |
|  | Internal | ADNI |  |  |  |  |
| **# Patients** | 5,645 | 1,737 | 321 | 837 | 1,756 | 293 |
| **Diagnosis** |  |  |  |  |  |  |
| MS | 806 | - | 137 | 138 | 263 | 63 |
| Non-MS | 4,839 | 1,737 | 184 | 699 | 1,493 | 230 |
| with WML | 2,660 | - | 76 | 326 | 461 | 113 |
| **Sex** |  |  |  |  |  |  |
| Women | 3,318 | 905 | 190 | 526 | 778 | 178 |
| Men | 2,327 | 830 | 131 | 311 | 802 | 115 |
| Unknown | - | 2 | - | - | 176 | - |
| **Age, years** | 52.1 ± 21.5 | 74.8 ± 8.4 | 44.5 ± 16.6 | 46.4 ± 17.1 | 52.7 ± 23.1<br>(N=1,686) | 45.3 ± 17.8 |
| **# Scans** | 6,261 | 1,857 | 332 | 837 | 1,756 | 293 |
| **MRI protocol** |  |  |  |  |  |  |
| 3D FLAIR | 1,622 | 1,456 | 293 | 630 | 1,052 | 293 |
| 2D FLAIR | 4,691 | - | 164 | 512 | 704 | - |
| 3D T1-w | 2,382 | 401 | 195 | 497 | 1,215 | 293 |
| 2D T1-w | 3,832 | - | 140 | 360 | 50 | - |
| 3D T1-CE | 1,384 | - | 189 | 414 | 784 | 293 |
| 2D T1-CE | 375 | - | 113 | 236 | - | - |
| Multi-Shell dMRI | 3,621 | - | 229 | 579 | - | - |
| **Field Strength** |  |  |  |  |  |  |
| 1.0 T | - | - | - | - | 4 | - |
| 1.5 T | 1 | 431 | - | - | 147 | 293 |
| 3.0 T | 6,260 | 1,423 | 332 | 837 | 1,499 | - |
| Unknown | - | 3 | - | - | 106 | - |
| **Scanner** |  |  |  |  |  |  |
| Siemens | 6,261 | 1,001 | 332 | 837 | 711 | - |
| GE | - | 685 | - | - | 594 | 293 |
| Philips | - | 167 | - | - | 326 | - |
| Others | - | - | - | - | 4 | - |
| Unknown | - | 4 | - | - | 121 | - |

**Table 2.**
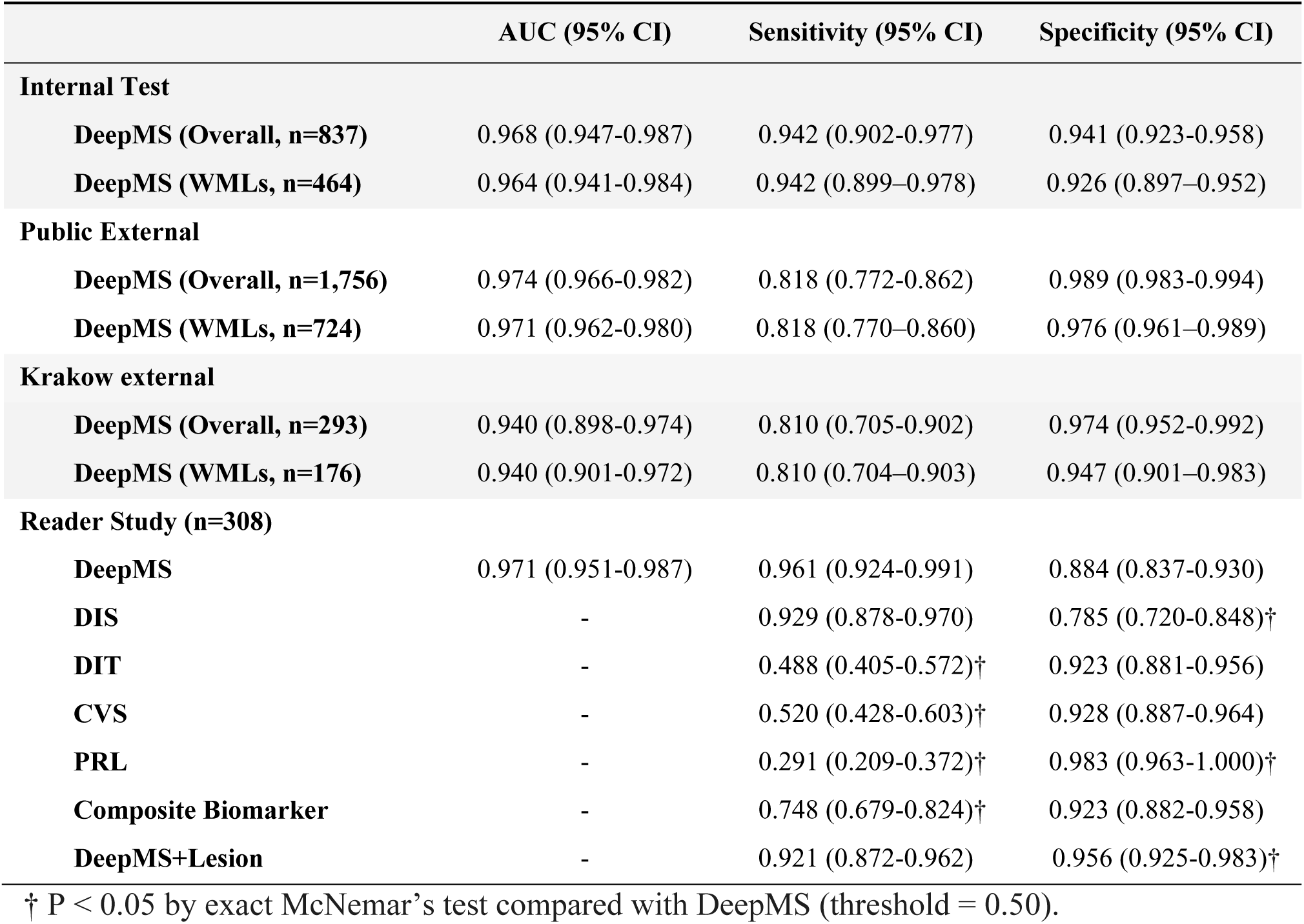
Performance metrics of the model and biomarkers.

To benchmark against the imaging biomarkers in the 2024 McDonald criteria^3^, we assembled a reader-study subgroup of WML-positive patients with available 3D FLAIR and susceptibility-weighted imaging. This enabled direct comparison between DeepMS and established MRI biomarkers—including DIS, DIT, CVS, PRL, and a composite biomarker defined according to the revised McDonald criteria (Fig. 1e). Finally, we performed ablation studies, lesion-masking experiments, and probability-map analyses to determine whether cross-modal co-training improved external generalisability, preserved diagnostic discrimination without visible WMLs, and shifted model activation into NAWM territories.

### Diagnostic performance across internal and external cohorts

DeepMS demonstrated highly robust diagnostic accuracy across all evaluation datasets (Table 2). In the internal test cohort (n=837; Fig. 2a), the model achieved an area under the receiver operating characteristic curve (ROC-AUC) of 0.968 (95% CI 0.947–0.987), maintaining an ROC-AUC of 0.964 (0.941–0.984) in the WML-positive subgroup (n=464). This performance generalised consistently to the Krakow cohort (n=293; Fig. 2b), with an overall ROC-AUC of 0.940 (0.898–0.974) and a WML-subgroup ROC-AUC of 0.940 (0.901–0.972). In the large public external cohort (n=1,756; Fig. 2c), overall ROC-AUC reached 0.974 (0.966–0.982), with the WML subgroup (n=724) at 0.971 (0.962–0.980). Sensitivity and specificity at the pre-specified operating point are summarised in Table 2. Dataset-specific performance within the public external cohort is provided in Supplementary Table 1. Performance stratified by age and sex is provided in Extended Data Fig. 2 and Supplementary Methods S11.

**Fig. 2:**
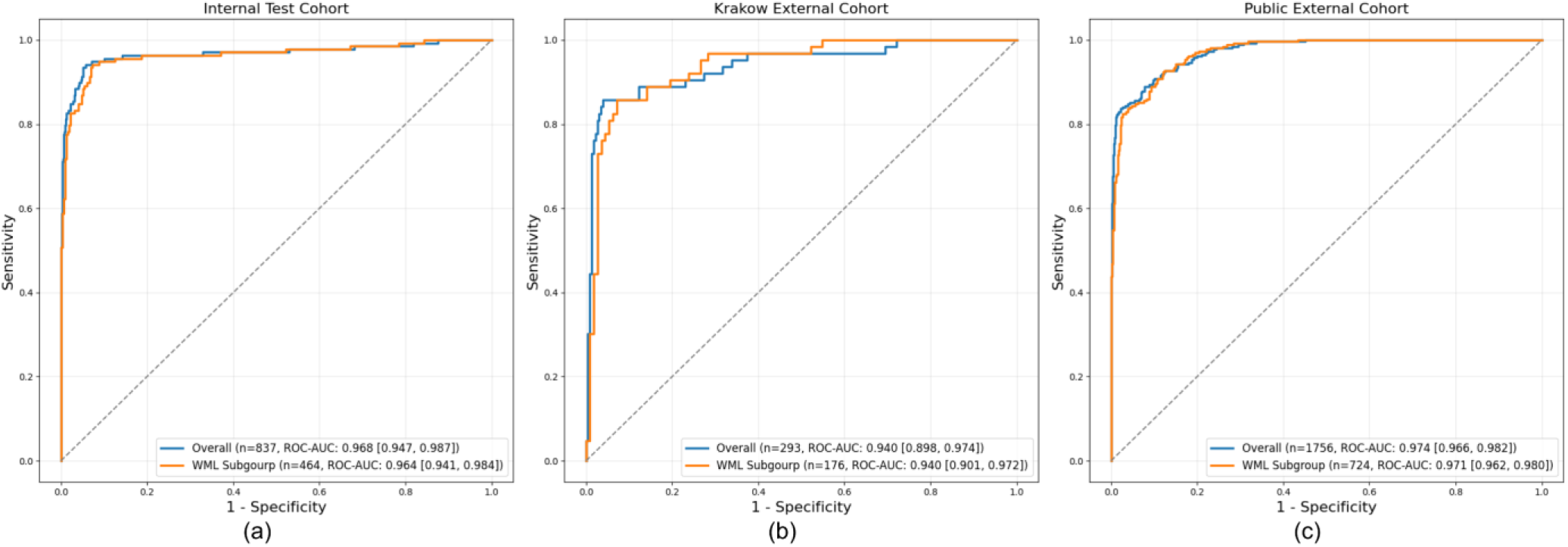
Diagnostic performance of DeepMS across internal and external cohorts. ROC curves for DeepMS in the internal test cohort (a), Krakow external cohort (b), and public external cohort (c), shown for the overall cohort and the WML subgroup. ROC-AUCs with 95% CIs are reported in the plot legends. WML, white matter lesion.

### Comparison with 2024 McDonald MRI biomarkers in the reader study

In the multi-reader study (n=308; Fig. 3a), DeepMS achieved an ROC-AUC of 0.971 (95% CI 0.951–0.987), with a sensitivity of 96.1% (92.4–99.1) and a specificity of 88.4% (83.7–93.0) (Table 2). By comparison, lesion-centric MRI biomarkers showed variable trade-offs between sensitivity and specificity: DIS, 92.9% (87.8–97.0) and 78.5% (72.0–84.8); DIT, 48.8% (40.5–57.2) and 92.3% (88.1–95.6); CVS, 52.0% (42.8–60.3) and 92.8% (88.7–96.4); PRL, 29.1% (20.9–37.2) and 98.3% (96.3–100.0); and the composite biomarker, 74.8% (67.9–82.4) and 92.3% (88.2–95.8), respectively (Table 2).

**Fig. 3:**
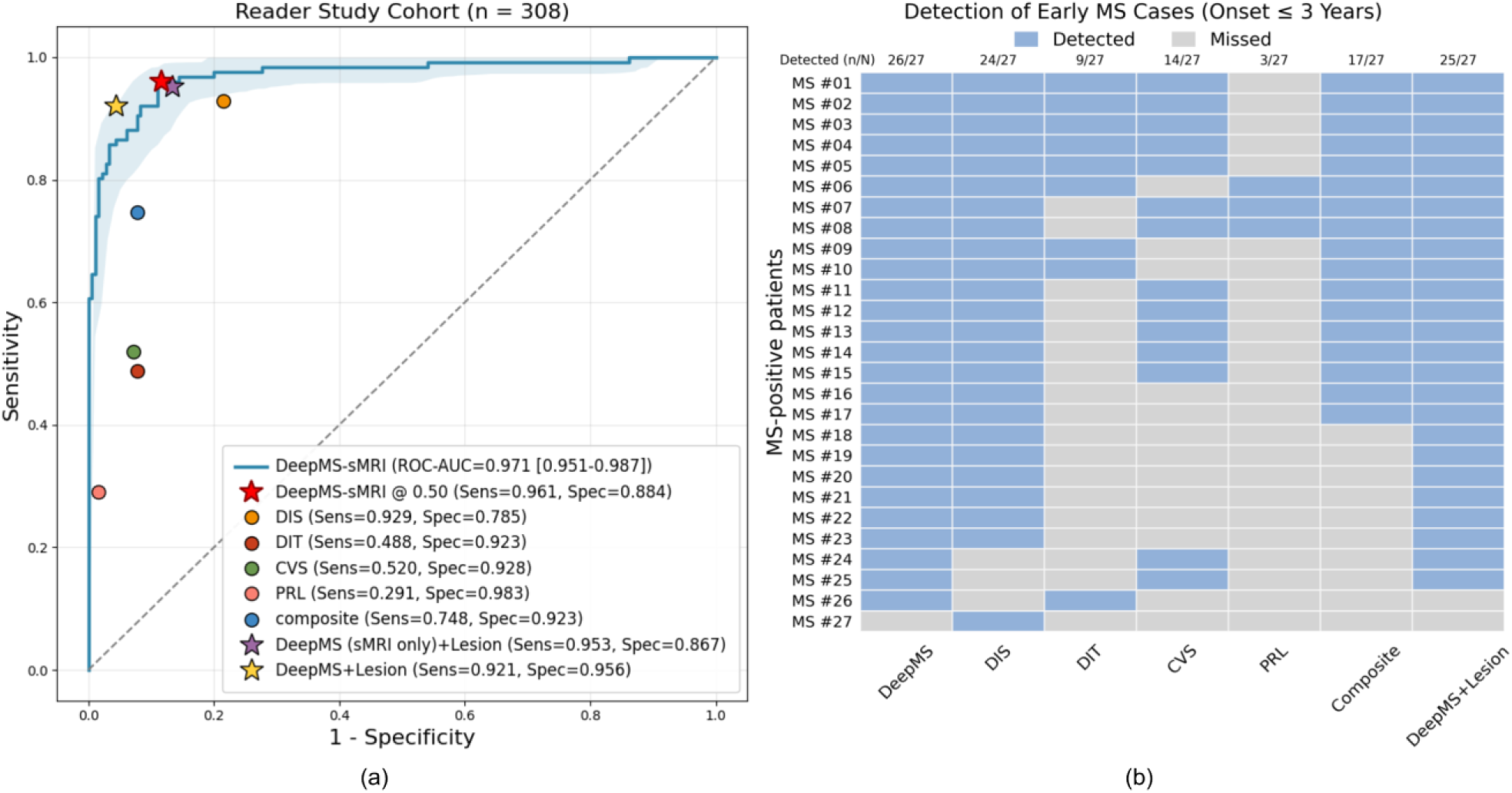
Benchmarking DeepMS against lesion-centric MRI biomarkers in the reader-study cohort and early-MS subgroup. **a,** In the reader-study cohort (n=308), DeepMS was compared with radiologist-assessed lesion-centric MRI biomarkers. The ROC curve and shaded area show the DeepMS performance and 95% CI, respectively; points show the sensitivity and specificity of individual biomarkers. Stars mark operating points for DeepMS at the prespecified threshold of 0.50 and for DeepMS+Lesion, a hybrid strategy requiring a positive DeepMS prediction plus at least one positive cross-sectional lesion biomarker: DIS, CVS, or PRL. **b,** Case-level detection in the early-MS subgroup, including 27 MS-positive patients with disease duration ≤3 years. Rows represent patients and columns represent DeepMS, lesion-centric biomarkers, the composite biomarker, and DeepMS+Lesion. Blue indicates detected cases and grey indicates missed cases; numbers above columns show the total detected cases for each method. CVS, central vein sign; DIS, dissemination in space; DIT, dissemination in time; MS, multiple sclerosis; PRL, paramagnetic rim lesion.

At matched operating points, DeepMS achieved a higher specificity than DIS (89.0% [84.4– 93.4] vs 78.5% [72.0–84.8]; p=0.006) while matching sensitivity; and a higher sensitivity than CVS (88.2% [82.1–93.1] vs 52.0% [42.8–60.3], p<0.001) and composite biomarker (90.6% [85.0–95.3] vs 74.8% [67.9–82.4], p=0.001) while matching specificity.

In early MS (defined as disease onset ≤ 3 years; n=27, Fig. 3b and Supplementary Table 2), DeepMS alone detected 26/27 cases, comparable to DIS (24/27; p=0.625) and outperforming CVS (14/27; p<0.001), PRL (3/27; p<0.001), and composite biomarker (17/27; p=0.008).

### Co-training improves external generalisability

Ablation studies showed that sMRI–dMRI co-training improved generalisability compared with training with sMRI only (Fig. 4a). In the FLAIR-only inference setting, this advantage was significant internally, where ROC-AUC improved from 0.945 (95% CI 0.916–0.970) to 0.963 (0.939–0.983) in the internal test cohort (DeLong’s test, p=0.020), and externally, where ROC- AUCs improved from 0.848 (0.785–0.901) to 0.944 (0.905–0.977) in the Krakow cohort (p<0.001) and from 0.922 (0.904–0.938) to 0.971 (0.961–0.979) in the public external cohort (p<0.001). Findings were consistent when all available sMRI sequences were used at inference, with a significant improvement in the public external cohort (Fig. 4b).

**Fig. 4:**
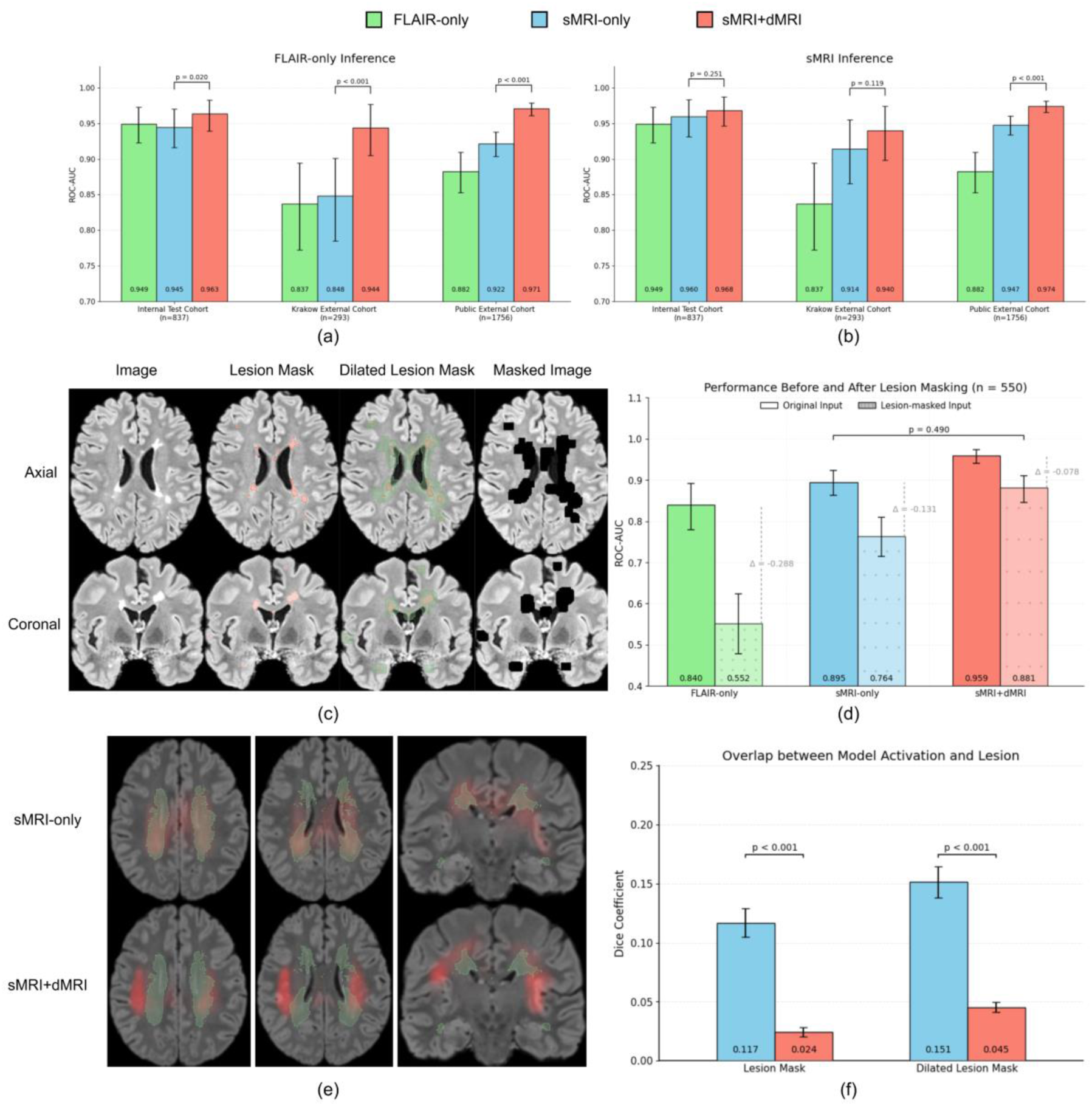
sMRI–dMRI co-training improves generalisability and captures NAWM-related diagnostic signals. **a,b,** ROC-AUCs of models trained with FLAIR only, sMRI only, or both sMRI and dMRI across the internal test cohort, Krakow external cohort, and public external cohort. Performance is shown under FLAIR-only inference (**a**) and routine sMRI inference using available sMRI from the same visit (**b**). **c,** Lesion-masking procedure. Lesion masks were spatially dilated to include lesional and perilesional regions, which were then digitally removed from the input images before inference. Red indicates the original lesion mask, and green indicates the dilated lesion mask. **d,** ROC-AUCs before and after lesion masking in the public external cohort subset with available lesion masks (n=550), stratified by training strategy. **e,** Group-level activation probability maps in external MS cases with available lesion masks (n=89). Red indicates model activation, and green indicates the lesion probability map. Compared with the sMRI-only model, the sMRI–dMRI co-trained model showed activation extending beyond focal lesion regions. **f,** Spatial overlap between model activation and lesion masks or dilated lesion masks, quantified using the mean Dice coefficient in external MS cases (n=89).

### Co-training preserves diagnostic discrimination after lesion removal

To determine the model’s dependence on WMLs, we performed digital lesion-masking in the subset of the public external cohort with available lesion masks (n=550). Lesional and perilesional voxels, defined using dilated lesion masks, were set to zero before inference (Fig. 4c), and model performance was compared before and after masking (Fig. 4d).

Models trained on sMRI alone showed substantial performance deterioration after lesion masking. ROC-AUC for the FLAIR-only model decreased from 0.840 (95% CI 0.780–0.892) to 0.552 (0.478–0.624; ΔROC-AUC −0.288), and that for the sMRI-only model decreased from 0.895 (0.863–0.923) to 0.764 (0.715–0.809; ΔROC-AUC −0.131). By contrast, the model co-trained with sMRI and dMRI was markedly more robust, with ROC-AUC decreasing from 0.959 (0.940–0.974) to 0.881 (0.846–0.911; ΔROC-AUC −0.078). After lesion masking, the co-trained model retained discrimination comparable to that of the sMRI-only model evaluated on the original unmasked input (0.881 versus 0.895; p=0.490).

### Co-training shifts model activation into NAWM regions

To characterise the spatial distribution of the model’s diagnostic signal, we aggregated activation heatmaps across MS cases with available lesion masks in the public external cohort (n=89) and generated group-level probability maps to characterise the spatial distribution of model activation (Fig. 4e). Qualitatively, DeepMS co-trained with both sMRI and dMRI showed activation extending beyond focal lesions into NAWM. In contrast, the corresponding sMRI- only-trained model showed activation concentrated predominantly within lesional and perilesional regions.

We next quantified the spatial overlap between model activation and lesion masks. Compared with the sMRI-only-trained model, the sMRI–dMRI co-trained DeepMS showed lower overlap with both lesion masks (mean Dice coefficient [mDice] 0.024 versus 0.117; p<0.001) and dilated lesion masks (0.045 versus 0.151; p<0.001) (Fig. 4f).

Unlike the sMRI-only model, in which NAWM activation decreased with increasing lesion load (Spearman’s r=−0.302, p=0.003; Extended Data Fig. 3a), the sMRI–dMRI co-training model showed no such inverse association (r=0.071, p=0.499; Extended Data Fig. 3b), suggesting that dMRI co-training prevented model activation from progressively collapsing into a purely lesion-centred pattern.

### DeepMS provides complementary diagnostic value to lesion-centric MRI biomarkers

To assess whether DeepMS provides complementary diagnostic value to established cross-sectional lesion-centric MRI biomarkers, we defined a simple hybrid strategy in which a case was classified as MS-positive only when DeepMS and at least one established cross-sectional biomarker (DIS, CVS, or PRL) were positive.

In the reader-study cohort, this hybrid approach achieved a sensitivity of 92.1% (95% CI 87.2– 96.2) and a specificity of 95.6% (92.5–98.3) (Fig. 3a; Table 2). Compared with the composite biomarker, the hybrid approach improved sensitivity (92.1% [87.2–96.2] versus 74.8% [67.9–82.4]; p<0.001), with no significant difference in specificity (95.6% [92.5–98.3] versus 92.3% [88.2–95.8]; p=0.210). In the early MS subgroup, the hybrid approach detected 25 of 27 cases, more sensitive than the composite biomarker (17 of 27; p=0.008; Fig. 3b). Compared to DeepMS alone, the hybrid approach significantly increased specificity (95.6% [92.5–98.3] versus 88.4% [83.7–93.0]; p<0.001). In contrast, the corresponding hybrid strategy based on the sMRI-only-trained model did not show the same complementary benefit, with performance remaining similar to that of the standalone model (Sensitivity 95.3% [91.3–98.5] versus 96.1% [92.4–99.1] and specificity 86.7% [81.7–91.1] versus 88.4% [83.7–93.0]). Its specificity was lower than the composite biomarker (86.7% [81.7–91.1] versus 92.3% [88.2–95.8]; p=0.041) (Fig. 3a).

### Representative clinical cases illustrate lesion-independent behaviour

Prediction heatmaps for representative clinical scenarios further highlight the mechanistic shift induced by dMRI co-training (Fig. 5). In a typical MS case (Fig. 5a), the sMRI-only-trained model focused predominantly on focal WMLs, whereas the co-trained model highlighted NAWM. In a complex MS mimic case that was clinically misdiagnosed as MS (meeting DIS criteria; Fig. 5b), the sMRI-only model yielded a false-positive classification based on lesion morphology. In contrast, the co-trained model correctly classified the case as negative. Failure cases were also observed, including a false-negative prediction in an MS case meeting DIS criteria (Fig. 5c).

**Fig. 5:**
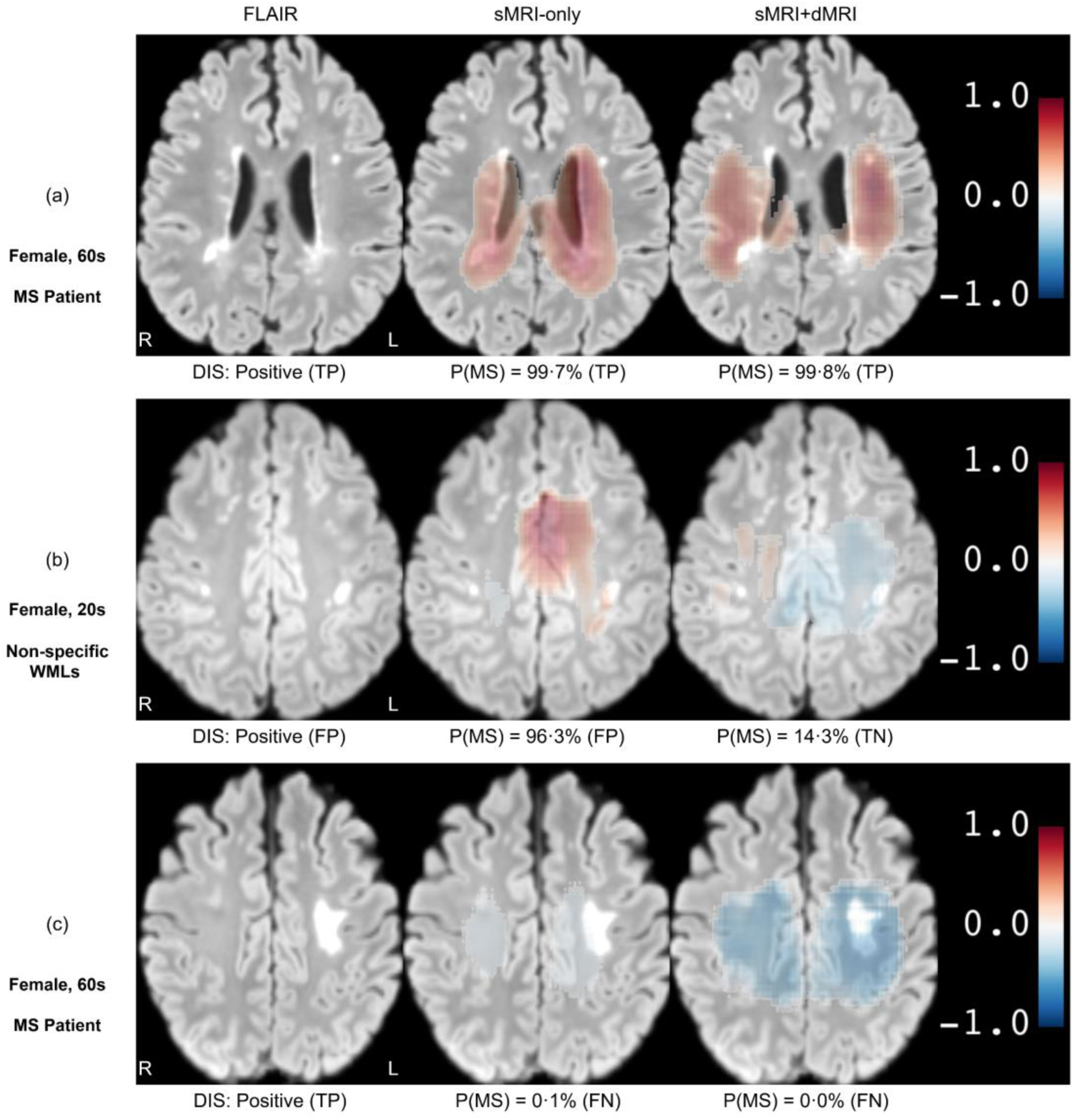
Prediction heatmaps across representative clinical scenarios on FLAIR MRI. Axial FLAIR images (left) are shown alongside prediction heatmaps from DeepMS trained on sMRI only (middle; sMRI-only) and DeepMS trained on both dMRI and sMRI (right; sMRI+dMRI). Red indicates local contributions supporting MS diagnosis, and blue indicates local contributions supporting non-MS diagnosis (scale −1 to 1); the predicted MS probability is shown below each heatmap. **a,** Typical MS case. Both models correctly identified MS, but compared with the sMRI-only model, the sMRI+dMRI co-trained model showed positive activation extending beyond focal lesions into NAWM. **b,** MS mimic with non-specific WMLs and DIS-positive imaging. The sMRI-only model yielded a false-positive prediction, whereas the sMRI+dMRI co-trained model correctly classified the case as non-MS, with broader negative activation in NAWM. **c,** False-negative MS case meeting DIS. Both models missed the case. DIS, dissemination in space; FP, false positive; FN, false negative; MS, multiple sclerosis; TN, true negative; TP, true positive.

Heatmaps visualised before and after lesion masking confirmed case-level stability (Extended Data Fig. 4). The co-trained model preserved its broad NAWM activation and prediction confidence despite the digital removal of lesions. Conversely, the sMRI-only model exhibited severe signal attenuation, spatial redistribution, and frequent diagnostic sign reversals once visible lesions were masked.

## Discussion

Current MS diagnosis remains centred on focal WMLs^3,17^, which are clinically important but inherently non-specific. On the other hand, while NAWM abnormalities contain microstructural pathology signals of MS^10,12^, they remain clinically underutilised as they are largely inaccessible on routine imaging. In this study, we addressed this gap by developing DeepMS, a cross-modal deep learning model trained on multi-shell dMRI and routine sMRI but designed to incorporate latent NAWM signatures using solely routine sMRI at deployment. Evaluated across diverse internal and external cohorts enriched with complex MS mimics, DeepMS showed robust performance, outperformed lesion-centred imaging biomarkers incorporated into the 2024 McDonald criteria^3^, and demonstrated complementary diagnostic value when combined with established lesion-centred biomarkers. These findings demonstrate that deep learning models can extract hidden NAWM signatures from routine scans, potentially advancing MS diagnosis beyond the current lesion-centric paradigm.

Compared with prior AI studies for MS diagnosis, our work advances the field in two aspects. First, existing AI systems have primarily focused on automating the detection and characterisation of visible WMLs^40–47^, rather than discovering complementary diagnostic signals beyond the conventional lesion-centric paradigm. In contrast, DeepMS was designed to recover diagnostically useful yet radiologically occult NAWM signatures and to test whether this hidden signal adds value beyond lesion-based assessment. In addition, earlier AI models were often developed for simplified classification tasks, such as distinguishing MS from healthy controls^45,46^ or a single mimic^40–42,47^, and rarely underwent broad external validation^40–46^ (Supplementary Table 8). In contrast, DeepMS was evaluated across 16 external datasets spanning clinically realistic populations with a broad spectrum of MS mimics, including small-vessel disease, migraine, and other demyelinating disorders. Importantly, we further benchmarked DeepMS against the imaging biomarkers incorporated into the 2024 McDonald criteria^3^, providing a clinically relevant reference point.

Established imaging biomarkers have substantially advanced MS diagnosis^3,5,6^, but they remain centred on visible lesions and therefore capture only a fraction of the disease^10,18^. This limitation is reflected in their characteristic trade-offs: DIS is highly sensitive yet prone to false positives in patients with non-specific WMLs^48,49^, whereas CVS^5^ and PRL^6^ offer higher specificity at the cost of lower sensitivity. DeepMS addresses this limitation by extracting NAWM-related signatures directly from routine sMRI and providing a complementary diagnostic dimension.

Adding this NAWM-related dimension to lesion-centred assessment increased sensitivity from 74.8% to 92.1% while maintaining high specificity compared with the composite of existing imaging biomarkers (95.6% versus 92.3%; p=0.210)^3^. More broadly, these findings suggest that MS diagnosis may benefit from evaluating both focal lesions and diffuse NAWM-related microstructural changes.

Several analyses support the interpretation that DeepMS complements established lesion-centred MRI biomarkers by capturing additional NAWM-related diagnostic information. In the lesion-masking experiment, DeepMS retained substantial discrimination performance after visible WMLs were removed, with performance comparable to the sMRI-only model evaluated on unmasked scans (ROC-AUC 0.881 versus 0.895, p=0.490). This finding indicates that it captured diagnostic signals beyond visible WMLs. In addition, activation mapping showed that, relative to sMRI-only training, sMRI–dMRI co-training shifted the model’s activation pattern away from focal WMLs and toward diffuse NAWM regions. Consistent with this observation, when combined with established lesion-centred biomarkers^3^, DeepMS provided added diagnostic value, whereas the sMRI-only comparator did not. Together, these findings suggest that the clinical gain provided by DeepMS is driven by extra-lesional information linked to diffuse NAWM pathology rather than by a reformulation of conventional lesion-based assessment.

This study has several limitations. First, the study was retrospective, and the effect of DeepMS on real-world diagnostic decision-making and patient outcomes remains to be tested prospectively. Second, DeepMS was developed primarily for brain MRI and does not yet incorporate other clinically important compartments in MS, particularly the spinal cord and optic nerve^3^. Third, although our hybrid strategy, which requires both DeepMS and at least one cross-sectional imaging biomarker to be positive, showed promising utility, this approach remains preliminary, and more principled fusion approaches should be explored. Finally, while multiple analyses suggest that DeepMS leveraged NAWM-related diagnostic information, this signal has not yet been directly validated histopathologically or distilled into a radiological pattern that clinicians can independently recognise and apply. Future work should focus on prospective validation, expanding to other anatomical regions, and converting the AI-derived NAWM signal into actionable biomarkers that physicians can easily interpret.

In conclusion, our work shows that deep learning models can recover diagnostically meaningful, non-lesional information from routine sMRI. By learning from paired multi-shell dMRI and sMRI data, DeepMS integrates focal lesion information with diffuse NAWM pathology to improve MS diagnosis. More broadly, our findings suggest that routine clinical imaging may contain latent disease signatures that are not visually apparent but can be extracted computationally. Unlocking these hidden signals through multimodal AI could not only advance MS diagnosis but also establish a broader framework for studying other neurological disorders where microscopic injury occurs before obvious structural abnormalities.

## Methods

### Study design

This HIPAA-compliant retrospective study aimed to develop and validate a deep learning model for diagnosing MS from routine sMRI. Fig. 1 provides an overview of the study workflow. The protocol was approved by the Institutional Review Board of NYU Grossman School of Medicine (IRB i14-01224, i25-01143), and the requirement for informed consent was waived. All analyses complied with the principles of the Declaration of Helsinki and were reported in accordance with the STARD 2015 guideline^50^.

### Study population and datasets

A cohort flowchart is provided in Extended Data Fig. 1. We retrospectively identified a consecutive cohort of clinically indicated brain MRI examinations acquired at an NYU Langone Health outpatient imaging centre (2014–2020). Eligible examinations were required to include single-/multi-shell dMRI and routine sMRI sequences as part of the clinical brain MRI protocol, yielding 7,493 studies from 6,825 patients. MS status was ascertained from the clinical record, based on diagnoses made by treating board-certified neurologists, according to standard diagnostic criteria^51,52^. We defined early MS as symptom onset ≤3 years before the index MRI, following prior work^53^. The internal dataset was split at the patient level into internal training, validation, and test sets. To increase diversity, training data were augmented with non-MS participants from the Alzheimer’s Disease Neuroimaging Initiative (ADNI)^24^.

For external validation, we included an independent cohort from the Clinical Imaging Diagnostics Center of the 5th Military Hospital with Polyclinic in Krakow (Krakow, Poland; 2020–2025). Inclusion criteria required complete 3D FLAIR, 3D T1-w, and 3D T1-CE sequences. MS diagnoses were established by neurologists according to the 2017 McDonald criteria^51^.

We further curated a cohort of 1,756 individuals from 15 publicly available datasets, covering healthy controls (MPI-Leipzig^28^), patients with MS (MSSEG31, MSSEG-232, OpenMS-Long36, OpenMS-Cross37, MS-ISBI27, PediMS38, QSM-MS33), and diverse non-MS neurological conditions, including demyelinating disease (PediDemi39), vascular WMH (WMH^35^, MRBrainS1834), stroke (ISLES-2022^29^), tumours (UCSF-PDGM25, MetsToBrain26), and epilepsy (OpenNeuro Epilepsy^30^). These datasets were collected across 13 international sites, and diagnostic labels were taken from the original dataset releases. Detailed cohort characteristics are provided in Extended Data Tables 1 and 2 and Supplementary Methods S1.

Uniform exclusion criteria were applied across all datasets, removing cases with clinically isolated syndrome (CIS) or radiologically isolated syndrome (RIS) without confirmed MS conversion, inadequate image quality, or missing FLAIR sequences. When two or more scans were available for the same patient in the test cohort, only the earliest scan was included. Cohort characteristics and exclusion flow are detailed in Table 1 and Extended Data Fig. 1.

### MRI acquisition and preprocessing

In the NYU Langone Health cohort, imaging was performed during routine clinical care on 3.0 T Siemens Magnetom Prisma or Skyra scanners. sMRI protocols included heterogeneous 2D and 3D acquisitions, comprising T1-w, T1-CE, and FLAIR. Susceptibility-weighted imaging (SWI) was acquired and used for the reader study to assess the CVS and PRL. dMRI analyses were restricted to multi-shell acquisitions obtained by a standard single-shot EPI protocol^54,55^. In the Krakow external cohort, only sMRI was used and was acquired on a 1.5 T GE SIGNA Artist system, including complete 3D T1-w, 3D T1-CE, and 3D FLAIR sequences. The public external cohort was restricted to sMRI and used heterogeneous protocols across vendors (Siemens, GE, Philips) and field strengths (predominantly 1.5–3 T). Detailed sequence parameters for all cohorts are provided in the Supplementary Methods S2 and Supplementary Tables 3-5.

All available raw sMRI data were processed using a pipeline comprising bias-field correction^56^, within-subject co-registration across sequences, skull stripping, and affine registration to MNI space. For dMRI, we employed the DESIGNER-v2 pipeline^54^ for denoising and artefact correction^57,58^ before estimating diffusion tensor (DTI)^59^, kurtosis (DKI)^19^, and standard model (SMI) imaging metrics^21^. Final model inputs were background-cropped, resized to 128 × 128 × 128 voxels, and scaled to the [0, 1] interval. Detailed algorithmic steps and software specifications are provided in Supplementary Methods S3-5 and Supplementary Figs. 1-2.

### DeepMS architecture

We developed DeepMS, a deep learning framework that predicts patient-level MS probability from at least one sMRI sequence (3D or 2D FLAIR, 3D T1-w, and 3D or 2D T1-CE) acquired during a single imaging visit. dMRI quantitative maps (DTI, DKI, and SMI) were used for training when available, but are not required for inference. The model uses a pretrained Swin- UNETR^23^ feature extractor and an attention-based multiple-instance learning (ABMIL) predictor^60^, with all parameters shared across MRI modalities. Each forward pass processes a single-modality 3D brain MRI volume and outputs a sequence-level MS logit (Extended Data Fig. 5).

### Model training

As shown in Fig. 1b, DeepMS was co-trained with both quantitative dMRI and sMRI to improve sensitivity to NAWM abnormalities. In the internal cohort, dMRI and sMRI were acquired during the same imaging session and were preprocessed to achieve approximate anatomical correspondence (Supplementary Methods S3). During training, each MRI sequence was treated as a separate input but passed through a unified, parameter-shared image encoder and supervised using the same patient-level diagnostic labels. Through this shared parameter optimisation, the model was encouraged to learn representations that were generalisable across modalities, enabling patterns learned from quantitative dMRI to inform the detection of subtle abnormalities on sMRI.

To reduce imbalance across disease labels and acquisition types, training used stratified oversampling over label-by-modality strata. DeepMS was optimised end-to-end using a composite objective that combined sequence-level diagnostic classification (ℒ_cls_) with spatial regularisation of model activation (ℒ_non-brain_ and ℒ_neg_):

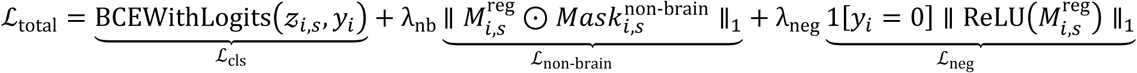

Here, 𝑧_𝑖,𝑠_ denotes the sequence-level logit for sequence 𝑠 from patient 𝑖, 𝑦_𝑖_ ∈ {0,1} denotes the patient-level diagnostic label, 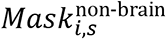 denotes the non-brain mask, ⊙ denotes element- wise multiplication, and 𝑀^reg^denotes a training-only auxiliary regularisation map, representing the spatial distribution of feature-level propensity toward the MS class before attention-based weighting (Extended Data Fig. 5a and Supplementary Methods S6–7). ℒ_cls_ optimised sequence- level diagnostic discrimination, ℒ_non-brain_ discouraged activation outside the brain, and ℒ_neg_ suppressed positive model activation in non-MS patients. λ_nb_ and λ_neg_ are hyperparameters that controlled the relative weights of the two regularisation terms.

Model checkpoints and hyperparameters were selected on the validation set using macro-averaged ROC-AUC across input modalities. Detailed training settings and hyperparameters are provided in Supplementary Methods S6–7 and Supplementary Table 6.

### Model inference and patient-level prediction

At inference (Fig. 1c), DeepMS operated on routine sMRI alone (2D or 3D FLAIR, 3D T1-w, and 2D or 3D T1-CE). Each available sMRI sequence was processed independently to produce a raw sequence-level logit. Before patient-level fusion, logits were calibrated by sequence-specific temperature scaling61, with all temperature parameters fitted exclusively on the internal validation set.

Patient-level predictions were generated using hierarchical logit fusion. Individual sequences were grouped into three predefined sequence types: FLAIR, T1-w, and T1-CE. For each patient, calibrated logits were first averaged within each available sequence type to prevent sequence types with more available sequences from dominating the patient-level prediction. The resulting sequence-type logits were then averaged across all available sequence types to obtain the patient-level logit:

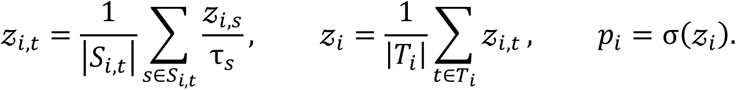

Here, 𝑖 indexes patients, 𝑠 indexes individual sMRI sequences, and 𝑡 indexes sequence types, with 𝑡 ∈ {FLAIR, T1-w, T1-CE}. 𝓏_𝑖,𝑠_ denotes the raw logit for sequence 𝑠 from patient 𝑖, τ_𝑠_ denotes the corresponding validation-fitted temperature parameter, 𝑆_𝑖,𝑡_ denotes the set of available sequences in type 𝑡 for patient 𝑖, and 𝑇_𝑖_ denotes the set of available sequence types for patient 𝑖. 𝑝_𝑖_ is the final patient-level MS probability. Detailed calibration and fusion procedures are provided in Supplementary Methods S8.

### Model interpretation with voxel-wise prediction heatmaps

To interpret sequence-level decisions, we generated voxel-wise prediction heatmaps, 𝑀^pred^, from the ABMIL interpretation branch (Extended Data Fig. 5a). For sequence 𝑠 from patient 𝑖, the heatmap  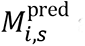 at voxel 𝑣 was defined as:

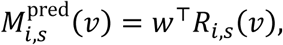

where 𝑅_𝑖,𝑠_(𝑣) is the attention-weighted representation at voxel 𝑣 and 𝑤 is the classifier weight vector. Each heatmap was divided by its maximum absolute activation, yielding a normalised heatmap 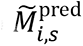 with value in [−1, 1], and was used only for visualisation and activation-based analyses. Detailed heatmap generation and normalisation procedures are provided in Supplementary Methods S6 and S9.

### Analysis of activation maps

For each sequence 𝑠 from the patient 𝑖, we derived an activation map 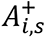 from the normalized prediction heatmap 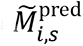 using a fixed heatmap threshold τ_*H*_ = 0.50:

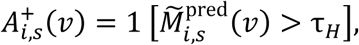

where 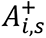 (*v*)denotes the binary activation at the voxel 𝑣.

Group-level activation-probability maps were generated by averaging binary activation maps voxel-wise across the external MS cases with available WML masks:

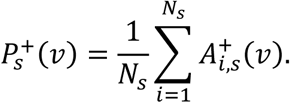

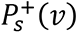 represents the proportion of MS cases with activation at the voxel 𝑣 in sequence 𝑠. Lesion probability maps were generated by averaging the binary WML masks across the same cases; voxels with lesion frequency greater than 10% were displayed in visualization (Fig. 4e). Detailed activation derivation and association analysis between lesion load and model activation are provided in Supplementary Methods S9.

### Reader study

To compare DeepMS with established MRI biomarkers, we conducted a reader study involving three board-certified neuroradiologists from NYU Langone Health (1-4 years post-fellowship experience; Fig. 1e) on a subset of the internal test cohort (n=308; 127 MS, 181 non-MS).

Inclusion required visible WMLs on 3D FLAIR, plus available SWI and a prior MRI (Extended Data Fig. 1). Scans were partitioned into three non-overlapping batches (93–109 each) and evaluated independently by blinded readers (age and sex provided). Following the 2024 McDonald criteria^3^, each reader assessed four MRI biomarkers: DIS, DIT, CVS (select-6 approach), and PRL, as part of their routine clinical image-reading protocol. MRI examinations were reviewed using Visage Imaging software, version 7.1.19.

Based on the McDonald criteria^3^, we defined a composite biomarker to integrate these four biomarkers. A case was classified as MS-positive if any condition was met: 1) DIS with DIT; 2) DIS with positive CVS; 3) DIS involving four characteristic regions, including periventricular, juxtacortical/cortical, infratentorial, and spinal cord regions; or 4) PRL with DIT and at least one characteristic region involved. Dedicated orbital imaging was not available, and optic nerve involvement was therefore not assessed. Cases not meeting these criteria were classified as composite-negative.

To evaluate whether DeepMS provided complementary information to current MRI biomarkers, we defined a hybrid strategy that required both a positive DeepMS prediction and at least one positive cross-sectional biomarker (DIS, CVS, or PRL) to classify a case as MS-positive.

### External generalisability and lesion-masking analyses

As shown in Fig. 4, we performed two analyses to evaluate the impact of dMRI-sMRI co-training. As T1-w and T1-CE sequences were not consistently available across public external datasets, we primarily evaluated on the FLAIR-only inference setting. First, to assess generalisability, we compared DeepMS with the corresponding sMRI-only trained model across the internal and external test cohorts. Second, to evaluate whether the model leveraged information from NAWM beyond WMLs, we conducted a lesion-masking experiment by comparing model performance on original versus digitally lesion-removed scans using a public external cohort subset with available WML masks (7 public datasets; n=550 [89 MS, 461 non-MS]). Lesion masks were dilated by three voxels (SciPy) and zeroed (ANTsPy) to remove visible lesion-related signal, with effectiveness confirmed by inspecting post-masking activations (Supplementary Fig. 3).

### Quantitative and statistical analysis

The primary performance metric was the area under the receiver operating characteristic curve (ROC-AUC). Secondary metrics were sensitivity and specificity, calculated at a decision threshold of 0.5 after calibration on the validation set (unless otherwise specified). Performance comparisons used DeLong’s test for ROC-AUC and exact McNemar’s test for paired sensitivity/specificity. In the reader study, DeepMS was further evaluated at operating points matched to the reference biomarkers: for DIS, we matched sensitivity to compare specificity; for CVS and the composite biomarker, we matched specificity to compare sensitivity. Associations between lesion burden and heatmap-derived activation volumes were assessed using Spearman’s rank correlation. All tests were two-sided, with p < 0.05 considered statistically significant.

Performance metrics were reported with 95% confidence intervals (CIs) estimated using 2,000 bootstrap resamples. Analyses were conducted in Python 3.11 (scikit-learn, SciPy).

## Data availability

The main data supporting the findings of this study are available within the Article, Extended Data, and Supplementary Information. Access links to all external public datasets used in this study, together with dataset-level documentation, are provided in the DeepMS GitHub repository. Individual-level data from the internal NYU Langone Health clinical cohorts are protected by patient privacy regulations and are not publicly available. Requests for access to deidentified internal data for non-commercial research purposes should be directed to the corresponding authors and will be reviewed by the NYU Langone Health Institutional Review Board and relevant institutional data-governance committees. Data access, if approved, will require a formal data-use agreement. The trained DeepMS model weights are not publicly available because they were developed using protected clinical data and are subject to institutional restrictions. Source data are provided with this paper.

## Code availability

The complete source code for model development, training, validation, and structural MRI (sMRI) preprocessing is publicly available at https://github.com/nyu-shenlab/DeepMS. The diffusion MRI (dMRI) preprocessing pipeline is available at https://github.com/NYU-DiffusionMRI/DESIGNER-v2.

## Data Availability

The source code for model development, training, validation, and structural MRI (sMRI) preprocessing is publicly available in the DeepMS repository (GitHub: nyu-shenlab/DeepMS). The diffusion MRI (dMRI) preprocessing pipeline is available at GitHub (NYU-DiffusionMRI/DESIGNER-v2). Links and identifiers for all external public datasets used in this study are provided in the DeepMS repository.
The internal clinical datasets and trained model weights are not publicly available due to patient privacy protections and institutional regulations. De-identified data access may be considered upon reasonable request, subject to review and approval by the Institutional Review Board of NYU Grossman School of Medicine / NYU Langone Health and execution of a Data Use Agreement.

https://github.com/nyu-shenlab/DeepMS

## Acknowledgments

This work was supported by the National Institute of Neurological Disorders and Stroke of the National Institutes of Health (NIH; R01 NS088040), the National Institute of Biomedical Imaging and Bioengineering of the NIH (R01 EB027075 and 1R01EB036530-01A1), and the Irma T. Hirschl Foundation. This work was performed at the Center for Advanced Imaging Innovation and Research (CAI2R), an NIBIB Biomedical Technology Resource Center (NIH P41 EB017183).

## Author Contributions

JM, EF, and YS conceived the study. JM, ML, VS and JC wrote the manuscript. JM, VS, WR, HCC, ML, AS, TP and JC did the data processing, experiment, and analysis. ML, IL, RP, MB, ML, TMS, KEH, and KO contributed materials and clinical expertise. DSN and EF contributed technical expertise on image processing and quantification. EF and YS supervised the work. All authors contributed to the experimental design, the interpretation of the results, and editing of the final manuscript. All authors had full access to the data, and JM, EF, and YS directly accessed and verified the data in the study. All authors accept the final responsibility to submit for publication and take responsibility for the contents of the manuscript.

## Competing interests

EF and DSN are co-inventors on patents and patent applications related to diffusion MRI, microstructural imaging, and MRI preprocessing. EF and DSN serve as advisory board members of Microstructure Imaging, Inc. and hold equity in the company. TMS is a founder of, holds equity in, and serves as a consultant for Microstructure Imaging, Inc. TMS also serves on the editorial board of Radiology. Microstructure Imaging, Inc. had no role in the study design, data collection, data analysis, data interpretation, manuscript preparation, or decision to submit the work for publication. The remaining authors declare no competing interests.

---

**Extended Data Fig. 1:**
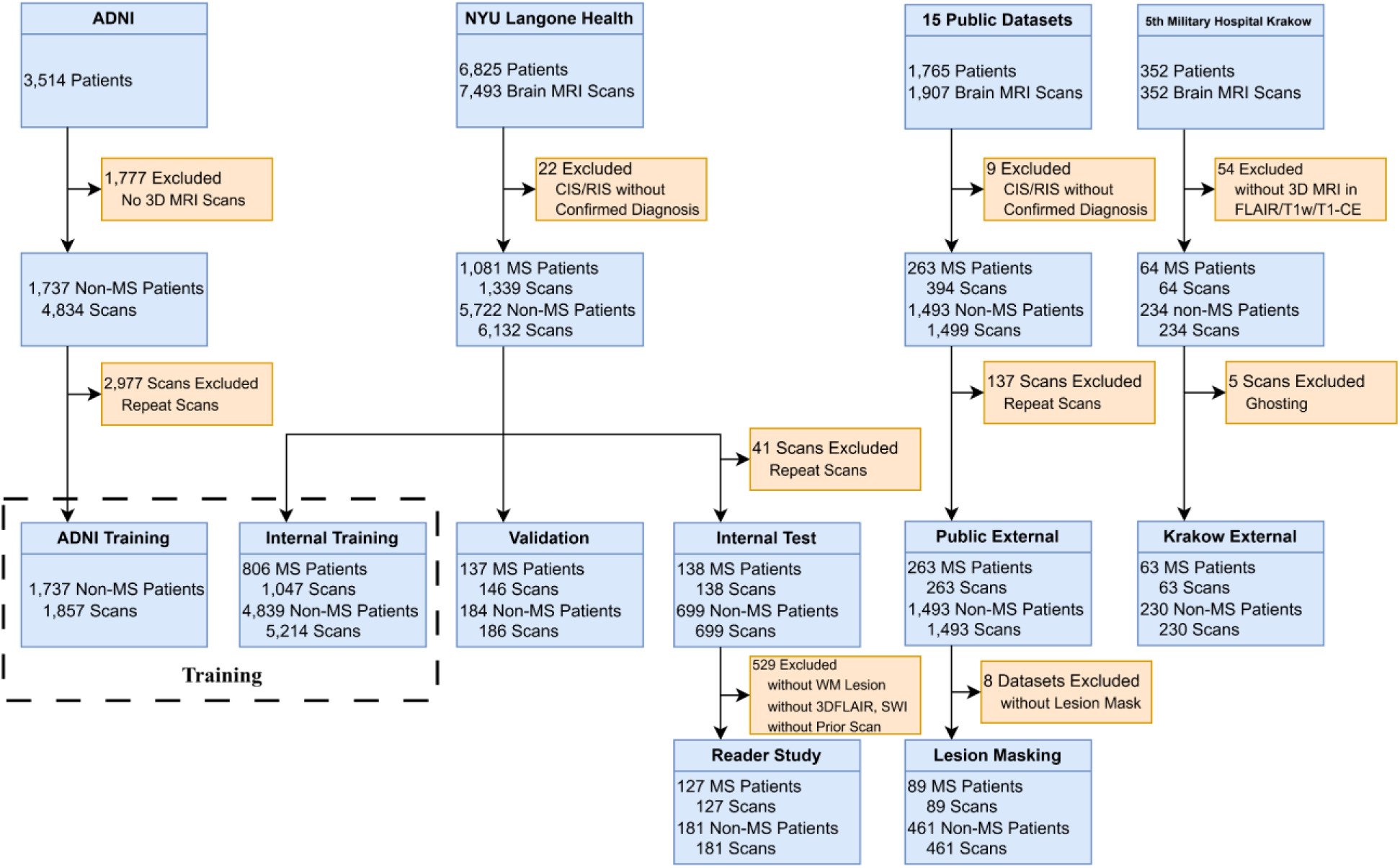
Study profile. Flowchart of data selection, exclusion criteria, and cohort assignment. A total of 10,589 patients with 11,336 brain MRI scans from NYU Langone Health, ADNI, the 5th Military Hospital with Polyclinic in Krakow, and 15 public datasets were processed. Patients with clinically isolated syndrome or radiologically isolated syndrome without a confirmed diagnosis of MS were excluded. The remaining data were assigned to the training set, validation set, internal test cohort, and two external test cohorts: the Krakow external cohort and public external cohort. In the test cohorts, only the first scan from each patient was retained. The reader study was performed in an internal-test subset with available baseline 3D FLAIR scans and white matter lesions. Lesion-masking experiments were performed in seven external datasets with available lesion masks. ADNI, Alzheimer’s Disease Neuroimaging Initiative; CIS, clinically isolated syndrome; RIS, radiologically isolated syndrome; WM, white matter.

**Extended Data Fig. 2:**
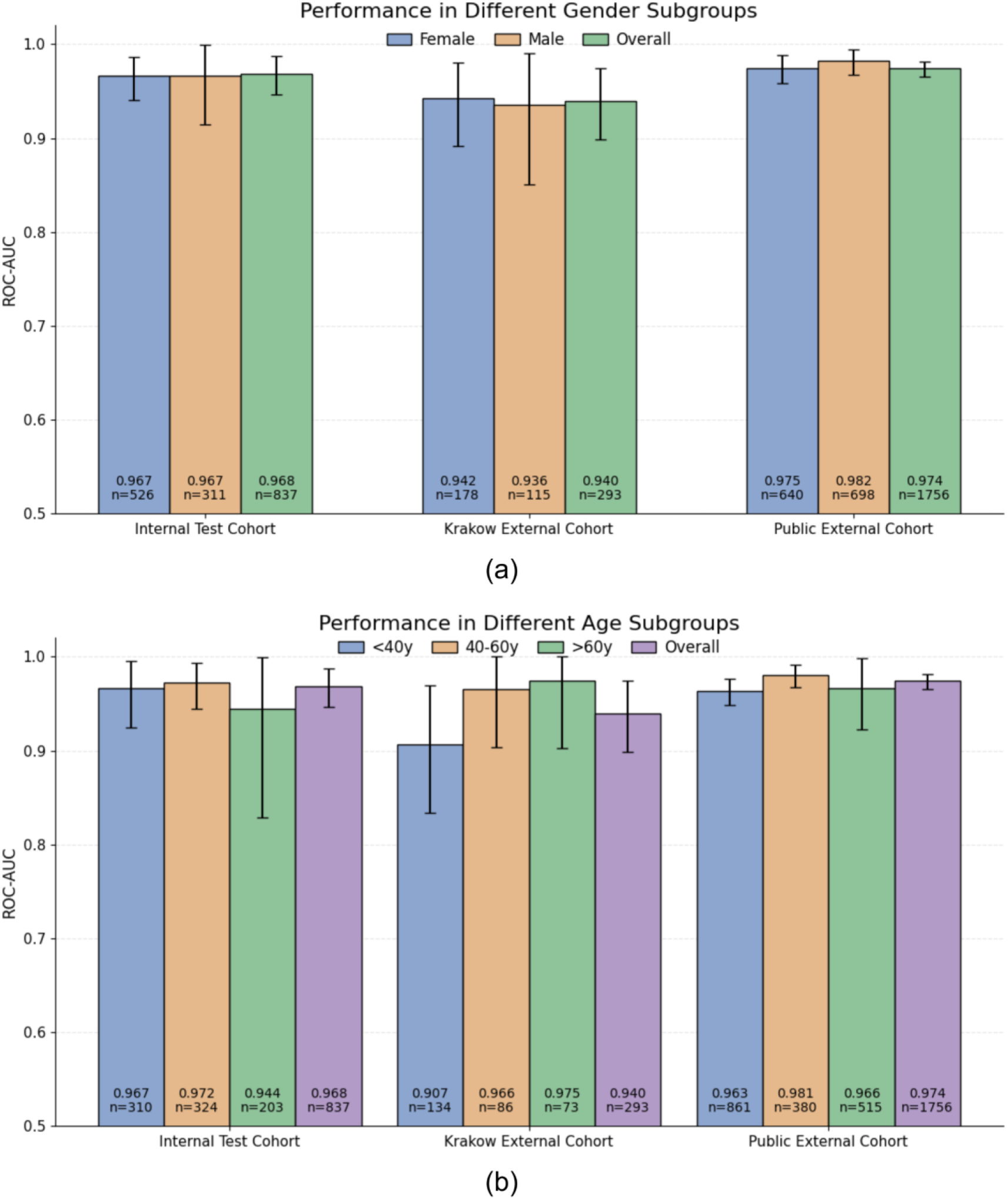
Subgroup performance of DeepMS across age and sex. **a,** ROC-AUCs of DeepMS in sex subgroups across the internal test cohort, Krakow external cohort, and public external cohort. Performance is shown for female and male patients, together with the overall cohort in each dataset. **b,** ROC-AUCs of DeepMS in age subgroups across the same cohorts. Performance is shown for patients aged <40 years, 40–60 years, and >60 years, together with the overall cohort in each dataset. Bars indicate ROC-AUCs with 95% CIs, and sample sizes are shown below the bars.

**Extended Data Fig. 3:**
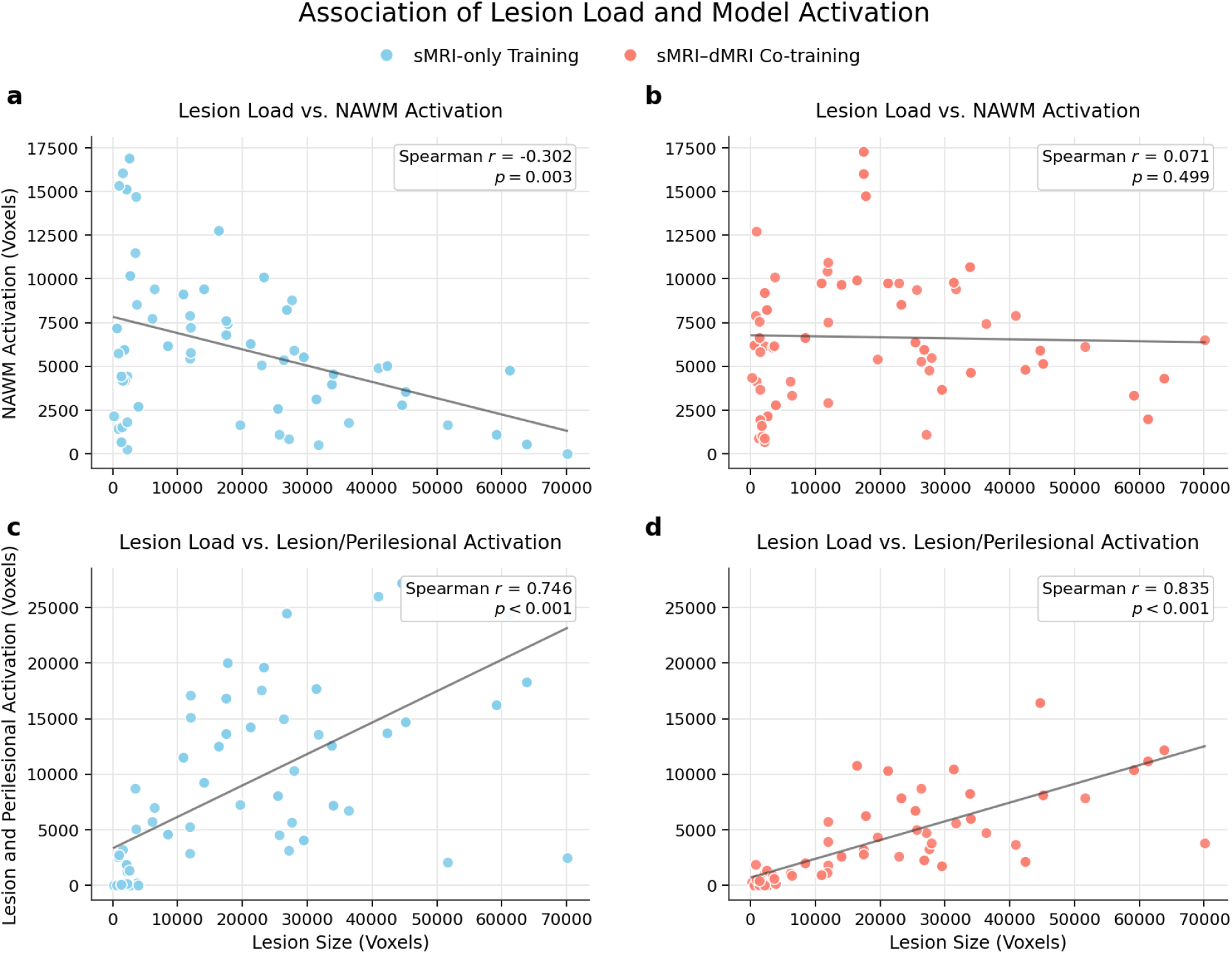
Association between lesion load and model activation under different training strategies. **a,b,** Relationship between lesion load and NAWM activation in the sMRI-only model (**a**) and the sMRI–dMRI co-trained model (**b**). The sMRI-only model showed an inverse association between lesion load and NAWM activation, whereas this association was absent after sMRI– dMRI co-training. **c,d,** Relationship between lesion load and lesion/perilesional activation in the sMRI-only model (**c**) and the sMRI–dMRI co-trained model (**d**). In both models, lesion load was positively associated with activation in lesion and perilesional regions. Lines indicate fitted trends, and inset values show Spearman’s correlation coefficients and two-sided P values. Axes represent voxel counts.

**Extended Data Fig. 4:**
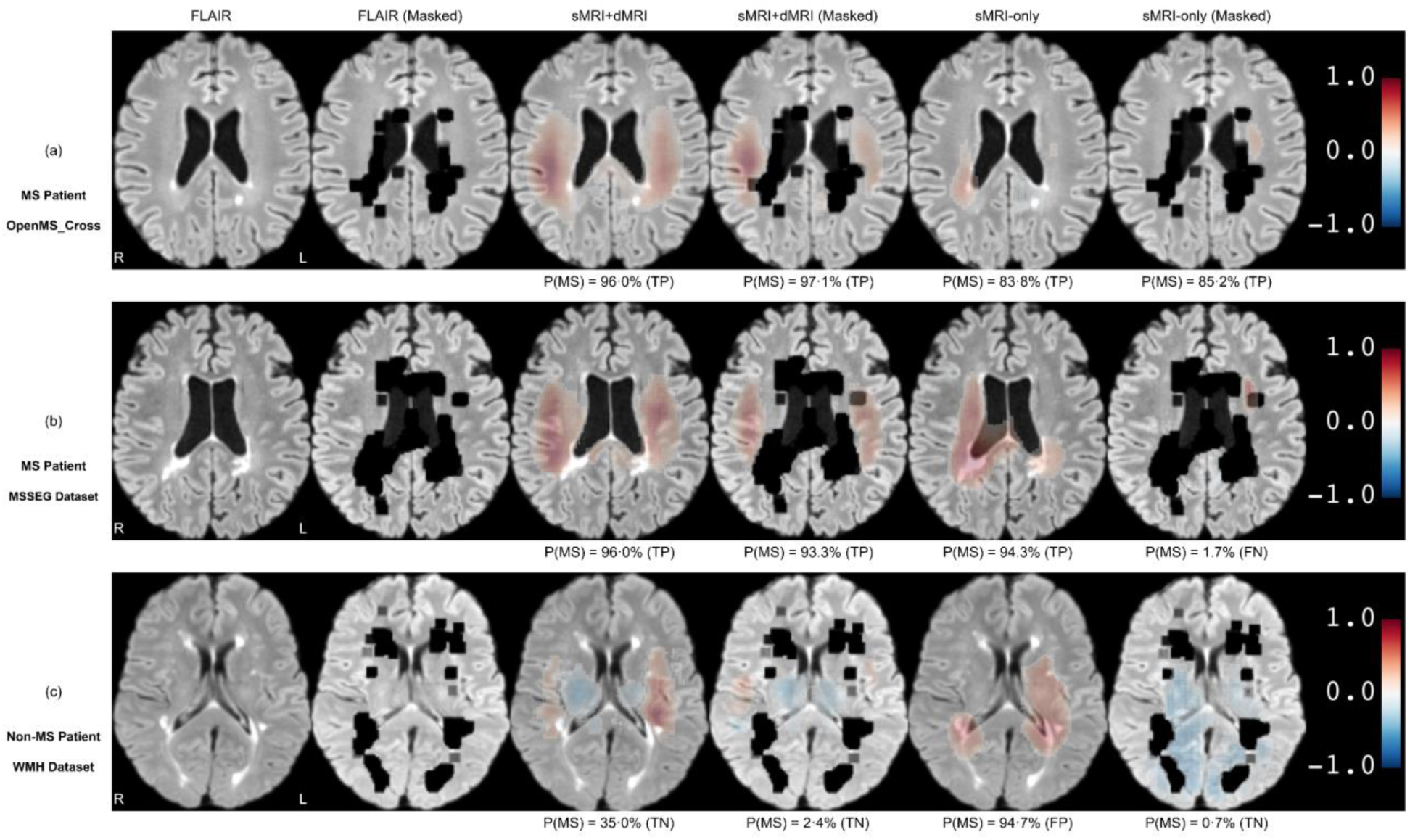
Case-level prediction heatmaps before and after lesion masking under different training settings. Representative axial FLAIR images and prediction heatmaps are shown for MS and non-MS cases before and after digital lesion masking. For each case, columns show the original FLAIR image, lesion-masked FLAIR image, heatmaps from the sMRI–dMRI co-trained model on original and masked inputs, and heatmaps from the sMRI-only model on original and masked inputs. Black regions indicate digitally masked lesions and perilesional voxels. Red indicates local contributions supporting MS classification, and blue indicates local contributions supporting non-MS classification (scale, −1 to 1). Predicted MS probabilities and outcome labels are shown below each heatmap. **a,b,** MS cases from the OpenMS-Cross and MSSEG datasets. The sMRI–dMRI co-trained model remained true positive after lesion masking, with persistent activation in NAWM regions, whereas the sMRI-only model showed attenuated or redistributed activation and, in one case, changed from true positive to false negative. **c,** Non-MS case from the WMH dataset. The sMRI-only model changed from false positive on the original image to true negative after lesion masking, whereas the sMRI–dMRI co-trained model remained true negative with qualitatively stable heatmaps. FN, false negative; FP, false positive; TN, true negative; TP, true positive.

**Extended Data Fig. 5:**
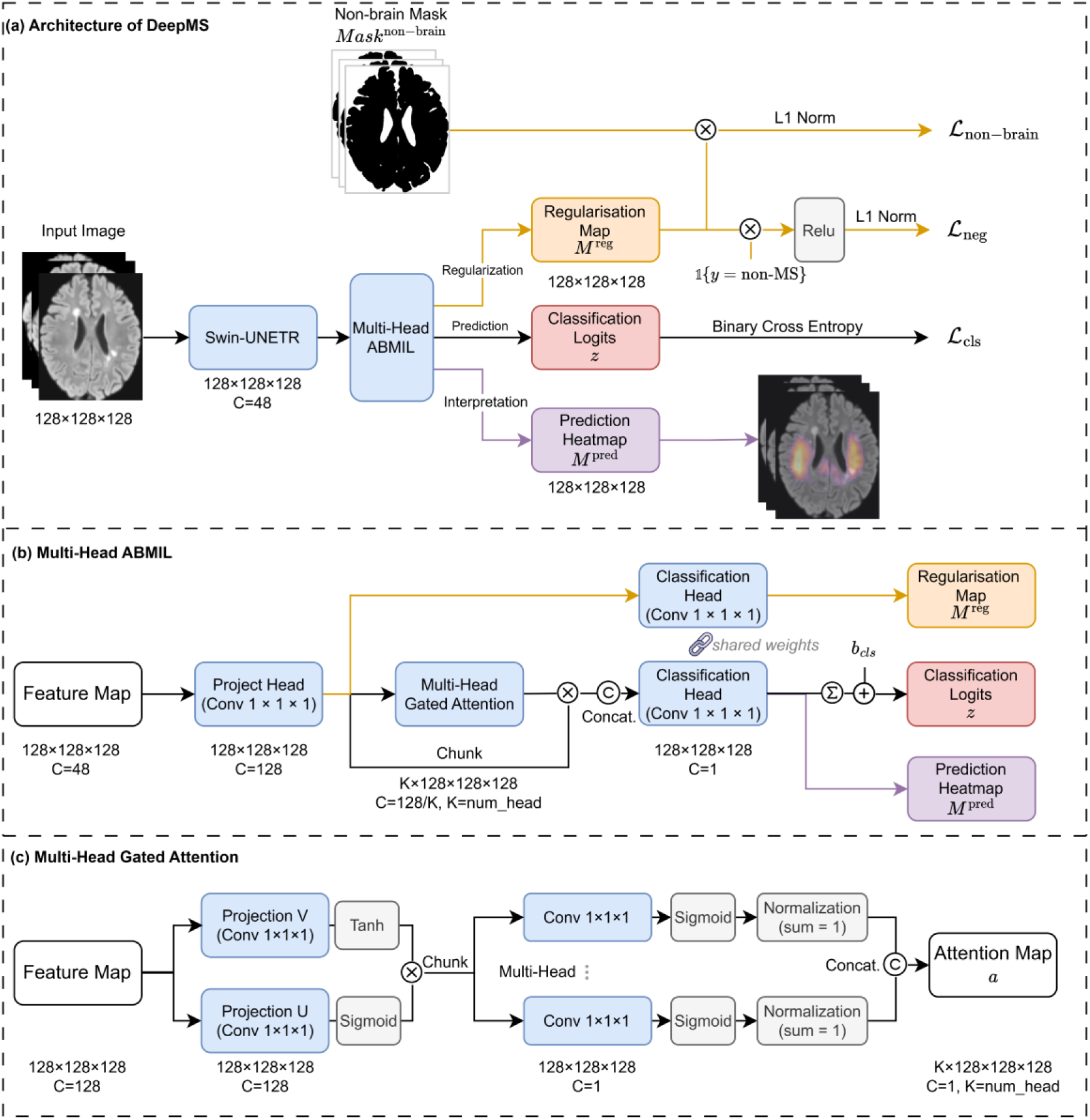
Architecture of the DeepMS framework. **a,** Overall DeepMS architecture. Each 3D MRI sequence, resampled to 128 × 128 × 128, is encoded by a VoComni-B-initialized Swin-UNETR backbone into a voxel-wise feature map (𝐶 = 48). A multi-head attention-based multiple-instance learning (ABMIL) module aggregates voxel features into a sequence-level diagnostic logit, 𝓏, supervised by the classification loss ℒ_cls_. During training, an auxiliary regularisation map, 𝑀^reg^, is used to penalise activation outside the brain through ℒ_non-brain_ and to suppress positive activation in non-MS cases through ℒ_neg_. An attention-weighted prediction heatmap, 𝑀^pred^, is generated for voxel-level interpretation. **b,** Multi-head ABMIL module. Backbone features are projected to a hidden space (𝐶 = 128) and assigned voxel-wise attention weights by the multi-head gated attention module. A shared 1 × 1 × 1 classification head is applied to unweighted features to produce 𝑀^reg^, and to attention-weighted features to produce 𝑀^pred^. The sequence-level logit 𝓏 is obtained by summing voxel-wise contributions and adding the classifier bias, 𝑏cls. **c,** Multi-head gated attention. Projected voxel features pass through tanh- and sigmoid-gated branches to form a gated representation. This representation is processed by 𝐾 = 2 parallel attention heads, normalised across voxels and concatenated to produce multi-head voxel-wise attention maps.

**Extended Data Table 1:** Summary of external test datasets (MS)

|  | MSSEG | MSSEG-2 | OpenMS-Long | OpenMS-Cross | MS-ISBI | PediMS | QSM |
| --- | --- | --- | --- | --- | --- | --- | --- |
| Institution (Country) | Inria (FR) | Inria (FR) | Univ. Ljubljana (SI) | Univ. Ljubljana (SI) | JHU (US) | Babeş-Bolyai Univ. (RO) | Univ. Bologna (IT) |
| # Patients | 53 | 40 | 15* | 27* | 19 | 9 | 150 |
| Sex |  |  |  |  |  |  |  |
| Women | 38 | NA | 13 | 21 | 15 | 7 | 89 |
| Men | 15 | NA | 2 | 6 | 4 | 2 | 61 |
| Age, years | 45.4 ± 10.3 | NA | 36.0 ± 10.2 | 40.1 ± 10.2 | 40.4 ± 9.2 | 13.9 ± 3.01 | 50.7 ± 14.1 |
| Diagnosis |  |  |  |  |  |  |  |
| MS | 53 | 40 | 15 | 27 | 19 | 9 | 100 |
| HC | — | — | — | — | — | — | 50 |
| # Scans | 53 | 40 | 15 | 27 | 19 | 9 | 150 |
| Modality |  |  |  |  |  |  |  |
| 3D FLAIR | 53 | 40 | 0 | 27 | 0 | 0 | 150 |
| 2D FLAIR | 0 | 0 | 15 | 0 | 19 | 9 | 0 |
| 3D T1-w | 53 | 0 | 0 | 27 | 19 | 0 | 150 |
| 3D T1-CE | 53 | 0 | 0 | 27 | 0 | 9 | 0 |
| Scanner |  |  |  |  |  |  |  |
| Siemens | 30 | 12 | 0 | 0 | 0 | 9 | 150 |
| GE | 8 | 0 | 0 | 0 | 0 | 0 | 0 |
| Philips | 15 | 28 | 15 | 27 | 19 | 0 | 0 |
| Others | 0 | 0 | 0 | 0 | 0 | 0 | 0 |
| Unknown | 0 | 0 | 0 | 0 | 0 | 0 | 0 |
| Field Strength |  |  |  |  |  |  |  |
| 1.0 T | 0 | 0 | 0 | 0 | 0 | 0 | 0 |
| 1.5 T | 15 | 9 | 0 | 0 | 0 | 0 | 0 |
| 3.0 T | 38 | 31 | 15 | 27 | 19 | 9 | 150 |
| Unknown | 0 | 0 | 0 | 0 | 0 | 0 | 0 |
| Lesion Mask | √ | × | × | √ | Partial | √ | × |

\* CIS patients without a confirmed diagnosis are excluded

**Extended Data Table 2:** Summary of external test datasets (non-MS)

|  | WMH | MRBrainS18 | ISLES-2022 | UCSF-PDGM | MetsToBrain | MPI-Leipzig | OpenNeuro epilepsy | PediDemi |
| --- | --- | --- | --- | --- | --- | --- | --- | --- |
| Institution (Country) | UMC Utrecht<br>NUHS Singapore<br>VU Amsterdam<br>(NL/SG) | UMC Utrecht<br>(NL) | Tech Univ.<br>Munich (DE) | UCSF (US) | Yale (US) | Max Planck<br>Inst. (DE) | Univ.<br>Hospital<br>Bonn (DE) | Babeş-Bolyai<br>Univ. (RO) |
| # Patients | 170 | 30 | 250 | 495 | 200 | 117 | 170 | 11* |
| Sex |  |  |  |  |  |  |  |  |
| Women | 85 | NA | 80 | 199 | 125 | 24 | 78 | 4 |
| Men | 85 | NA | 64 | 296 | 75 | 93 | 92 | 7 |
| Age, years | 70.1 ± 9.3 | NA | 72.3 ± 13.0 | 56.9 ± 15.1 | 63.8 ± 11.8 | 36.0 ± 18.1 | 28.9 ± 12.4 | 8.9 ± 4.7 |
| Diagnosis |  |  |  |  |  |  |  |  |
| MS | — | — | — | — | — | — | — | — |
| WML | 170 | 30 | — | — | — | — | — | — |
| Tumor | — | — | — | 495 | 200 | — | — | — |
| CVD | — | — | 250 | — | — | — | — | — |
| Epilepsy | — | — | — | — | — | — | 85 | — |
| Demyelin | — | — | — | — | — | — | — | 11 |
| HC | — | — | — | — | — | 117 | 85 | — |
| # Scans | 170 | 30 | 250 | 495 | 200 | 117 | 170 | 11 |
| Modality |  |  |  |  |  |  |  |  |
| 3D FLAIR | 0 | 0 | 0 | 495 | 0 | 117 | 170 | 0 |
| 2D FLAIR | 170 | 30 | 250 | 0 | 200 | 0 | 0 | 11 |
| 3D T1-w | 170 | 30 | 0 | 495 | 0 | 117 | 170 | 11 |
| 3D T1-CE | 0 | 0 | 0 | 495 | 200 | 0 | 0 | 0 |
| Scanner |  |  |  |  |  |  |  |  |
| Siemens | 50 | 0 | 4 | 0 | 158 | 117 | 170 | 11 |
| GE | 60 | 0 | 0 | 495 | 31 | 0 | 0 | 0 |
| Philips | 60 | 30 | 125 | 0 | 7 | 0 | 0 | 0 |
| Others | 0 | 0 | 0 | 0 | 4 | 0 | 0 | 0 |
| Unknown | 0 | 0 | 121 | 0 | 0 | 0 | 0 | 0 |
| Field Strength |  |  |  |  |  |  |  |  |
| 1.0 T | 0 | 0 | 0 | 0 | 4 | 0 | 0 | 0 |
| 1.5 T | 10 | 0 | 0 | 0 | 113 | 0 | 0 | 0 |
| 3.0 T | 160 | 30 | 144 | 495 | 83 | 117 | 170 | 11 |
| Unknown | 0 | 0 | 106 | 0 | 0 | 0 | 0 | 0 |
| Lesion Mask | √ | √ | √ | Tumour | Tumour | × | × | √ |

785 \* CIS patients without a confirmed diagnosis are excluded

## Notes

### Funding Statement

This work was supported by the National Institute of Neurological Disorders and Stroke, National Institutes of Health (NIH) (R01 NS088040); the National Institute of Biomedical Imaging and Bioengineering, NIH (R01 EB027075 and 1R01EB036530-01A1); the NIBIB Biomedical Technology Resource Center, NIH (P41 EB017183); and the Irma T. Hirschl Foundation.

### Author Declarations

Institutional Review Board of New York University Grossman School of Medicine gave ethical approval for this work and waived the requirement for informed consent (IRB i14-01224; i25-01143).

### Summary of Updates

This revised version has been substantially updated to improve the clarity, focus, and completeness of the manuscript. First, we revised the writing and overall framing to place greater emphasis on the central biological and clinical question: whether diagnostic information related to normal appearing white matter can be recovered from routine structural MRI using cross-modal deep learning. The introduction, results, and discussion were reorganized and streamlined to make this narrative clearer. Second, we added additional analyses to further support the interpretation that the model uses information beyond visible white matter lesions. These include expanded activation analyses, lesion masking experiments, quantitative assessment of activation overlap with lesion and perilesion regions, and additional comparisons across internal and external cohorts. These revisions provide stronger evidence that DeepMS captures complementary normal appearing white matter-related signals rather than relying only on focal lesion burden. Third, we expanded the visual and methodological documentation. We added new and revised figures to better summarize the model architecture, cohort design, diagnostic performance, interpretability analyses, and external validation results. We also substantially expanded the Methods and Supplementary Methods to provide clearer descriptions of preprocessing, model training, inference, calibration, activation mapping, lesion masking, statistical analysis, and sensitivity analyses.

