## Supplementary Material for "Diagnosis of Multiple Sclerosis Using Multimodal Deep Learning Integrating Lesion and Normal-Appearing White Matter: A Retrospective Study with International Multicentre External Validation"

S1. Public external cohort details	2

S2. MRI acquisition	4

S3. Structural MRI preprocessing	5

S4. Diffusion MRI preprocessing and diffusion parameter estimation	6

S5. Input transformation and augmentation	7

S6. Model architecture	8

S7. Model training	10

S8. Model inference	12

S9. Interpretation analysis	14

S10. Calibration analysis	17

S11. Subgroup analyses	18

Supplementary Fig. 1 | Structural MRI preprocessing pipeline.	19

Supplementary Fig. 2 | Data preprocessing and augmentation workflows.	20

Supplementary Fig. 3 | Effect of lesion masking on model activation in lesion and perilesional regions.	21

Supplementary Fig. 4 | Calibration curves of the DeepMS model across internal and external cohorts.	22

Supplementary Table 1 | Model performance in each public external dataset	23

Supplementary Table 2 | Sensitivity analysis of DeepMS versus biomarkers in early MS (≤3 years)	24

Supplementary Table 3 | Structural MRI acquisition parameters in internal cohort	25

Supplementary Table 4 | Diffusion MRI acquisition parameters in internal cohort	26

Supplementary Table 5 | Structural MRI acquisition parameters in Krakow external cohort	27

Supplementary Table 6 | Summary of implementation details and hyperparameters.	28

Supplementary Table 7 | Calibration analysis of DeepMS in different cohorts	29

Supplementary Table 8 | Summary of prior MRI-based AI studies for multiple sclerosis diagnosis	30

STARD 2015 Checklist	32

Supplementary Reference	34

### S1. Public external cohort details

We assembled a public external cohort comprising 15 public datasets to assess model generalisability and robustness. All analyses were conducted at the patient level. When multiple MRI examinations were available for an individual, we retained the earliest eligible scan to prevent double-counting. Public external-cohort inference used structural MRI (sMRI) only; diffusion MRI (dMRI), quantitative susceptibility mapping (QSM), and other advanced modalities were not used as model inputs. Individuals with clinically isolated syndrome (CIS) or radiologically isolated syndrome (RIS) without a confirmed subsequent MS diagnosis were excluded.

#### 1. Public MS cohorts

**MSSEG (n=53; Inria, France)**^1^**.** This public benchmark comprises adults with MS and multi-sequence structural MRI. MS lesion masks were delineated by seven expert raters and aggregated into a consensus reference.

**MSSEG-2 (n=40; Inria, France)**^2^**.** This public benchmark comprises longitudinal MS imaging with paired 3D FLAIR at two timepoints acquired across multiple sites and scanner vendors, with consensus annotations for new lesions between timepoints.

**OpenMS-Long (n=15; University of Ljubljana, Slovenia)**^3^**.** This cohort comprises adults with MS from a longitudinal study with baseline and follow-up multi-sequence structural MRI. In the original release (20 participants), five individuals labelled as CIS without a subsequently confirmed diagnosis were excluded, consistent with our study-wide exclusion criteria.

**OpenMS-Cross (n=27; University of Ljubljana, Slovenia)**^4^**.** This cohort comprises a cross-sectional sample of adults with MS and multi-sequence structural MRI (including 3D FLAIR). Reference white-matter lesion masks were generated via a multi-rater consensus process involving three expert raters. Of the 30 participants in the original cohort, we excluded three individuals labelled as CIS without a subsequently confirmed diagnosis, consistent with our study-wide exclusion criteria.

**MS-ISBI (n=19; Johns Hopkins University, USA)**^5^**.** This cohort comprises adults with clinically definite MS enrolled in a longitudinal imaging study with multi-sequence structural MRI. Reference manual lesion masks were provided for the original training subset only (5/19 participants).

**PediMS (n=9; Babeș-Bolyai University, Romania)**^6^**.** This cohort comprises longitudinal brain MRI from nine children with relapsing–remitting MS (onset age 6–17 years), with 1–6 examinations per individual. Structural MRI sequences (including FLAIR and T1-weighted imaging) were available; expert MS lesion segmentations were provided in the source dataset.

**QSM (n=150; University of Bologna, Italy)**^7^**.** This single-centre cohort includes 100 adults with MS and 50 healthy controls; MS was diagnosed per the 2017 McDonald criteria. MRI was acquired using a multiparametric protocol that included structural MRI (T1-weighted and FLAIR) and an acquisition for QSM reconstruction.

#### 2. Public MS mimic cohorts

**WMH (n=170; multi-site: Utrecht/Singapore/Amsterdam)**^8^**.** This cohort comprises memory-clinic participants with ageing-related vascular and degenerative pathology, enriched for white-matter hyperintensities of presumed vascular origin (cerebral small-vessel disease).

**MRBrainS18 (n=30; UMC Utrecht, Netherlands)**^9^**.** This cohort comprises adults aged >50 years with vascular and neurodegenerative comorbidities and matched controls at increased cardiovascular risk; brain atrophy and white-matter lesions are common and may resemble MS-associated hyperintensities on FLAIR.

**PediDemi (n=11; Babeș-Bolyai University, Romania)**^10^**.** This paediatric cohort comprises children with non-MS demyelinating disorders (e.g., ADEM and NMOSD spectrum disorders) and was included as MS differential diagnoses in paediatric presentations. Of the 13 participants in the original release, we excluded two individuals labelled as CIS without a subsequently confirmed diagnosis, consistent with our study-wide exclusion criteria.

#### 3. Other public neurological disease cohorts

**ISLES-2022 (n=250; Technical University of Munich, Germany)**^11^**.** This cohort comprised adults undergoing clinical brain MRI for suspected or confirmed acute-to-subacute ischaemic stroke.

**UCSF-PDGM (n=495; UCSF, USA)**^12^**.** Adults with histopathologically confirmed WHO grade II–IV diffuse gliomas who underwent pre-operative MRI and initial tumour resection at a single tertiary centre (2015–2021); individuals with prior brain-tumour treatment were excluded.

**MetsToBrain (n=200; Yale University, USA)**^13^**.** This cohort comprises patients with clinically or pathologically confirmed brain metastases identified from Yale New Haven Hospital registries (2013–2021), with pre-treatment MRI meeting the required structural sequences and quality criteria.

**OpenNeuro epilepsy (n=170; University Hospital Bonn, Germany)**^14^**.** This cohort comprises 85 individuals undergoing pre-surgical evaluation for focal epilepsy due to focal cortical dysplasia type II (FCD II) and 85 healthy controls.

#### 4. Public healthy control cohort

**MPI-Leipzig (n=117; Max Planck Institute, Germany)**^15^**.** This cohort comprises community-recruited healthy younger and older adults who underwent structured medical screening; individuals with a history of neurological disorders (including multiple sclerosis, stroke, or epilepsy) were excluded. We used participants with available high-resolution structural MRI in this study.

### S2. MRI acquisition

#### 1. Internal MRI acquisition

All internal MRI examinations were performed on 3.0 T Siemens Magnetom Prisma or Skyra scanners as part of routine clinical brain imaging. Structural and diffusion MRI protocols were standardised across scanners, with minor scanner-level variation in acquisition parameters (Supplementary Tables 3 and 4).

##### **1.1 Structural MRI**

The structural protocol included 3D T1-weighted (T1-w) MPRAGE with or without contrast, 3D FLAIR, 2D FLAIR, 2D T1-w with or without contrast, and susceptibility-weighted imaging (SWI) (Supplementary Table 3).

3D T1-w was acquired with TR ≈ 2100 ms, TE ≈ 2.7 ms, TI ≈ 900 ms, flip angle 8°, and 1.0 × 1.0 × 1.0 mm³ isotropic voxels, using the same protocol for contrast-enhanced and non-contrast scans. 3D FLAIR was acquired with TR ≈ 6000 ms, TE ≈ 325 ms, TI ≈ 2100 ms, flip angle 120°, and 1.0 × 1.0 × 1.0 mm³ isotropic voxels.

When 3D acquisitions were not available, 2D T1-w and 2D FLAIR sequences with in-plane resolution 0.9–1.0 × 0.9–1.0 mm² and 3.0 mm slice thickness were used. SWI was acquired with similar spatial resolution (0.9–1.0 × 0.9–1.0 × 3.0 mm³) and was used only for qualitative assessment in the neuroradiologist reader study; SWI images were not used as inputs to the deep-learning model.

##### **1.2 Diffusion MRI**

Diffusion MRI was performed using single-shot spin-echo echo-planar imaging (EPI) sequences (Supplementary Table 4). A multi-shell protocol was used with 5 b = 0 s/mm² images, 4 diffusion directions at b = 250 s/mm², 20 directions at b = 1000 s/mm², and 60 directions at b = 2000 s/mm². Typical parameters were TR 3.2–4.3 s, TE 70–100 ms, voxel size 1.7 × 1.7 × 3.0 mm³ (50 slices), partial Fourier 6/8, and a total acquisition time of approximately 7 minutes. One additional b = 0 s/mm² volume with reversed phase-encoding polarity was acquired for EPI distortion correction using standard methods^16,17^. In this study, only this multi-shell diffusion protocol was used to derive diffusion metrics; single-shell clinical diffusion sequences were not used.

#### 2. Krakow cohort MRI acquisition

All MRI examinations in the Krakow external cohort were acquired on 1.5 T GE SIGNA Artist scanners as part of routine clinical brain imaging. Only structural MRI was available for this cohort (Supplementary Table 5).

##### **2.1 Structural MRI**

The structural protocol included 3D T1-w (with and without contrast) and 3D FLAIR (Supplementary Table 5). 3D T1-w (± contrast) was acquired with TR ≈ 8.5 ms, TE ≈ 3.6 ms, TI ≈ 450 ms, and flip angle ≈ 12°, with voxel size ranging from ≈0.4–0.6 × 0.4–0.6 × 1.0–1.6 mm³. 3D FLAIR was acquired with TR ≈ 6200 ms, TE ≈ 90 ms, TI ≈ 1760 ms, and flip angle ≈ 90°, with voxel size ranging from ≈0.4–0.6 × 0.4–0.6 × 1.0–1.6 mm³.

### S3. Structural MRI preprocessing

All available raw structural MRI (sMRI) volumes from the internal cohort and, when available, from external cohorts were processed using a standardised preprocessing pipeline (Supplementary Fig. 1), implemented primarily with ANTsPy^18^ and HD-BET^19^. For each patient, all structural sequences, including 3D/2D FLAIR and 3D/2D T1-w or T1-CE, were processed jointly.

First, each volume underwent N4 bias-field correction^20^ and intensity clipping (0.1–99.9th percentile), followed by reorientation to a standard left–posterior–inferior (LPI) orientation.

Second, within-subject multi-sequence co-registration was performed in native (subject) space. For each patient, a reference structural sequence was selected using a fixed hierarchy that prioritised 3D FLAIR, followed by 3D T1-w, 2D T1-w, 2D FLAIR, and other sequences. All remaining structural images were affinely registered to this reference using ANTsPy.

Third, brain extraction was performed on the reference image using HD-BET, yielding a skull-stripped reference image and a subject-specific brain mask. This mask was subsequently applied to all co-registered structural sequences to remove non-brain tissue in subject space. The resulting co-registered, skull-stripped images in native space were used as inputs for model training.

Fourth, for validation and inference, the skull-stripped reference image was additionally affinely registered to the MNI ICBM152 2009a symmetric template using ANTs registration. The resulting transform was applied to all skull-stripped, co-registered structural sequences, resampling them to 1 × 1 × 1 mm³ in MNI space across modalities. A final intensity-clipping step (0–99.9th percentile) was applied in template space to harmonise dynamic range across subjects and sites; these MNI-normalised images were used as inference inputs.

For external datasets where only pre-processed sMRI volumes were available (for example, already skull-stripped and/or registered in the parent study), we did not reapply the full pipeline. Instead, images were reoriented and, when needed, affinely registered to the MNI template.

### S4. Diffusion MRI preprocessing and diffusion parameter estimation

We applied the DESIGNER (Diffusion parameter EStImation with Gibbs and NoisE Removal) preprocessing pipeline^21,22^ to all multi-shell dMRI datasets to remove noise and image artefacts. The pipeline consisted of:

(a) MP-PCA (Marchenko–Pastur principal component analysis) adaptive-patch denoising^23,24^ with eigenvalue shrinkage^25^;
 (b) removal of partial-Fourier–induced Gibbs ringing (RPG)^26^;
 (c) echo-planar imaging (EPI) distortion correction using a reversed phase-encoding b = 0 image;
 (d) eddy-current and motion correction^27^;
 (e) b = 0 intensity normalisation; and
 (f) Rician bias correction^28^.

After preprocessing, we used the DESIGNER tissue microstructure imaging (TMI) module for diffusion parameter estimation. We derived the following sets of diffusion metrics:

- **DTI (diffusion tensor imaging)**^29^: mean diffusivity (MD), radial diffusivity (RD), axial diffusivity (AD), and fractional anisotropy (FA).
- **DKI (diffusion kurtosis imaging)**^30,31^: mean kurtosis (MK), radial kurtosis (RK), and axial kurtosis (AK).
- **SMI (standard model imaging of white matter)**^32–34^: extra-axonal diffusivity parallel (De∥) and perpendicular (De⟂) to the axon, intra-axonal axial diffusivity (Da), axonal water fraction *f* (IAS / (IAS + EAS)), and *p*₂, the anisotropy invariant of the fibre ODF.

Before being used as model input, all diffusion-derived quantitative maps were intensity-normalised using fixed, physiologically motivated scaling. For each metric, values were first clipped to a plausible range and then divided by a predefined upper bound so that the final maps lay approximately in [0, 1]. Specifically, fractional anisotropy (FA), axonal water fraction *f* and *p*₂ were clipped to [0, 1]; diffusivity-type measures (MD, RD, AD from tensor/kurtosis models, as well as Da, De∥ and De⟂ from SMI) and mean/axial kurtosis (MK, AK) were clipped to [0, 3] and scaled by 1/3; and radial kurtosis (RK) was clipped to [0, 5] and scaled by 1/5. This global normalisation yielded comparable dynamic ranges across diffusion-derived inputs while preserving the relative contrast between different metrics.

### S5. Input transformation and augmentation

#### 1. Data transformation

All MRI volumes were processed using a unified 3D preprocessing pipeline implemented in Python with MONAI (v1.4.0) and PyTorch. To maintain voxel-wise correspondence, all geometric operations were applied identically to each image volume and any associated binary masks (e.g., lesion masks), when available.

The input-transformation workflow (Supplementary Fig. 2) included reorientation to a canonical right-anterior-superior (RAS) coordinate system, intensity-based foreground cropping to remove background air, and resampling to a fixed grid of 128 × 128 × 128 voxels. NaN values were set to zero. Intensity normalisation was modality-specific: sMRI volumes were min-max scaled to [0,1], whereas dMRI parametric maps were scaled to [0,1] using a pre-specified upper bound. Non-brain masks were derived during preprocessing and used in downstream analyses.

#### 2. Data augmentation

On-the-fly augmentation was applied during training to improve robustness to acquisition-related variability (Supplementary Fig. 2).

Spatial augmentations were applied identically to images and any available masks, including random axis flips (probability 0.5) and random affine transformations (probability 0.5; rotations up to 5° per axis; translations up to 5 voxels).

Intensity-based augmentations were applied to images only, including random gamma adjustment (0.7–1.3; probability 0.3), bias-field simulation (probability 0.3), additive Gaussian noise (probability 0.3), and Gaussian smoothing (probability 0.3).

### S6. Model architecture

#### 1. Overview

DeepMS consists of a volumetric feature extractor and an attention-based multiple-instance learning (ABMIL) predictor^35^. Each forward pass processes one preprocessed 3D MRI volume, denoted $X_{i,s}$ for sequence $s$ from patient $i$, and outputs a raw sequence-level MS logit $\mathcal{z}_{i,s}$. The same network parameters were shared across all eligible sMRI sequences and diffusion-derived inputs used during training. Input transformation and augmentation are described in Supplementary Methods S5.

#### 2. Feature extractor

We used Swin-UNETR as the backbone^36^, initialised with the VoComni-B self-supervised foundation-model weights^37^. Following the VoComni-B configuration, inputs were processed through the full encoder-decoder pathway, and decoder features were used for downstream modelling to preserve spatial granularity. The backbone maps input $X_{i,s}\in\mathbb{R}^{1\times128\times128\times128}$ to a spatially congruent feature map $F_{i,s}\in\mathbb{R}^{C\times128\times128\times128},$ where $C=48$. A $1\times1\times1$ convolution then projects these features to $D_{\text{hidden}}=128$ channels. The projected voxel-wise feature vector at voxel $v\in\Omega_{i,s}$ is denoted $h_{i,s}\left( v \right)\in\mathbb{R}^{D_{\text{hidden}}}$.

#### 3. Multi-head ABMIL predictor

The ABMIL predictor treats each MRI volume as a bag and voxel-wise features as instances. A gated attention representation was computed for each voxel as:

$$\mathbf{g}_{i,s}(v)=\sigma\left( \mathbf{W}_{U}\mathbf{h}_{i,s}(v)+\mathbf{b}_{U} \right)\odot\tanh\left( \mathbf{W}_{V}\mathbf{h}_{i,s}(v)+\mathbf{b}_{V} \right),$$

where $W_{U}$ and $W_{V}$are $1\times1\times1$ convolutions, $b_{U}$ and $b_{V}$ are learnable biases, $\sigma$ denotes the sigmoid function, and $\odot$ denotes element-wise multiplication. Dropout with probability 0.25 was applied during training.

We used $K=2$ attention heads. For head $k$, a grouped convolution produced a raw attention score $\mathcal{l}_{i,s,k}(v)$ from $g_{i,s}\left( v \right)$. Scores were normalised with sigmoid-based attention:

$$\alpha_{i,s,k}(v)=\frac{\sigma\left( \mathcal{l}_{i,s,k}(v) \right)}{\sum_{u\in\Omega_{i,s}} \sigma\left( \mathcal{l}_{i,s,k}(u) \right)}, k=1,\ldots,K.$$

The projected features were split into head-specific partitions $h_{i,s,k}\left( v \right)\in\mathbb{R}^{D_{\text{hidden}}/K}.$ The attention-weighted voxel representation used by the classifier was defined as:

$\mathbf{R}_{i,s}(v)=\mathrm{Concat}\left( \alpha_{i,s,1}(v)\mathbf{h}_{i,s,1}(v),\ldots,\alpha_{i,s,K}(v)\mathbf{h}_{i,s,K}(v) \right),$

where $R_{i,s}\left( v \right)\in\mathbb{R}^{D_{\text{hidden}}}$. The global sequence representation was obtained by summing the attention-weighted representations over voxels:

$$r_{i,s}=\sum_{v\in\Omega_{i,s}} R_{i,s}\left( v \right)$$

#### 4. Classification branch

A linear classifier was implemented as a $1\times1\times1$convolution. Let $w\in\mathbb{R}^{D_{\text{hidden}}}$ denote the classifier weight vector and $b$ denote the learnable classifier bias. The sequence-level logit was computed as:

$$\mathcal{z}_{i,s}=w^{\top}r_{i,s}+b.$$

#### 5. Voxel-wise maps for regularisation and interpretation

The same classifier weights were used to derive two voxel-wise maps with distinct roles (Extended Data Fig. 5). First, the training-only auxiliary regularisation map was computed on unweighted voxel features:

$$M_{i,s}^{\text{reg}}\left( v \right)=w^{\top}h_{i,s}\left( v \right).$$

$M_{i,s}^{\text{reg}}$ was used only for the non-brain and negative-case regularisation terms described in Supplementary Methods S7.

Second, the voxel-wise prediction heatmap was computed on the attention-weighted voxel representation:

$$M_{i,s}^{\text{pred}}\left( v \right)=w^{\top}R_{i,s}\left( v \right).$$

This heatmap provides a voxel-level decomposition of the sequence-level decision before the classifier bias, because:

$$\sum_{v\in\Omega_{i,s}} M_{i,s}^{\text{pred}}\left( v \right)=w^{\top}\sum_{v\in\Omega_{i,s}} R_{i,s}\left( v \right)=\mathcal{z}_{i,s}-b.$$

### S7. Model training

#### 1. Training data and inputs

DeepMS was trained on a composite multimodal dataset combining the internal training cohort and the ADNI database. Each training instance consisted of a single preprocessed 3D MRI volume $X_{i,s}$ of fixed spatial size $128\times128\times128$ and the corresponding patient-level label $y_{i}\in\{0,1\}$. Training inputs included sMRI sequences (2D/3D FLAIR, 2D/3D T1-w, and 2D/3D T1-CE), the diffusion $b=0$ image, and DTI-, DKI-, and SMI-derived quantitative maps.

#### 2. Model training framework

In the internal cohort, same-session dMRI and sMRI scans were foreground-cropped and resampled to a common grid to establish approximate anatomical correspondence. All modalities were processed by the same image encoder and ABMIL predictor. This parameter-sharing strategy encouraged representations that generalise across modalities, allowing microstructural information learned from dMRI-derived maps to inform the detection of subtle abnormalities on routine sMRI.

#### 3. Sampling strategy

To address the imbalance in both disease labels and imaging modalities, we used stratified oversampling. Strata were defined as the Cartesian product of disease label and input modality. At each optimisation step, one non-empty stratum was sampled with equal probability, followed by uniform sampling with replacement of one 3D volume from that stratum. This prevented parameter updates from being dominated by the most prevalent label or acquisition protocol.

#### 4. Loss functions and training objective

The network was trained end-to-end using a composite objective that optimised sequence-level classification while regularising the training-only map $M_{i,s}^{\text{reg}}$defined in Supplementary Methods S6:

$$\mathcal{L}_{\text{total}}=\underset{\mathcal{L}_{\text{cls}}}{\underbrace{\text{BCEWithLogits}\left( \mathcal{z}_{i,s},y_{i} \right)}}+\underset{\mathcal{L}_{\text{non-brain}}}{\lambda_{\text{nb}}\underbrace{\parallel M_{i,s}^{\text{reg}}\odot Mask_{i,s}^{\text{non-brain}}\parallel_{1}}}+\underset{\mathcal{L}_{\text{neg}}}{\lambda_{\text{neg}}\underbrace{\boldsymbol{1}\left[ y_{i}=0 \right]\parallel\text{ReLU}\left( M_{i,s}^{\text{reg}} \right)\parallel_{1}}}$$

Here, $\mathcal{z}_{i,s}$ is the raw sequence-level logit, $y_{i}$ is the patient-level diagnostic label, $Mask_{i,s}^{\text{non-brain}}$ is the binary non-brain mask for the input volume, and $1\left[ \cdot\right]$ is the indicator function. $\mathcal{L}_{\text{cls}}$optimised diagnostic discrimination, $\mathcal{L}_{\text{non-brain}}$ discouraged activation outside the brain, and $\mathcal{L}_{\text{neg}}$ suppressed positive model activation in non-MS cases.

We set $\lambda_{\text{nb}}={10}^{-6}$. For $\lambda_{\text{neg}}$, we used a warm-up schedule: $\lambda_{\text{neg}}=0$ for epochs 1-4, $\lambda_{\text{neg}}$ increased linearly from ${10}^{-8}$ at epoch 5 to 1$0^{-7}$at epoch 15; and $\lambda_{\text{neg}}=0$ thereafter.

#### 5. Optimisation, implementation, and model selection

Models were implemented in PyTorch (v2.6.0) and trained on two NVIDIA A100 GPUs using automatic mixed precision (FP16). Optimisation used AdamW $(\beta_{1}=0.9$, $\beta_{2}=0.99$9) with a global batch size of 16, an initial learning rate of ${10}^{-4}$, and a cosine learning-rate schedule. Training proceeded for up to 40 epochs, with early stopping if the model-selection metric did not improve for five consecutive epochs.

After each epoch, checkpoints were evaluated on the validation set. The final checkpoint was selected using macro-averaged ROC-AUC, computed as the unweighted mean ROC-AUC across input modalities, to prioritise cross-modality generalisation within the shared-parameter model. Implementation details and hyperparameters are summarised in Supplementary Table 6.

### S8. Model inference

#### 1. Eligible sMRI inputs

At inference, DeepMS processed routine sMRI only. Each available sequence was processed independently to produce a raw sequence-level logit $\mathcal{z}_{i,s}$; all dMRI-derived maps and other non-sMRI sequences were excluded. Eligible sequence types followed the 2021 MAGNIMS-CMSC-NAIMS brain MRI consensus protocol^38^ and comprised:

- **FLAIR:** 2D FLAIR, 3D FLAIR
- **T1-w:** 3D T1-w
- **T1-CE:** 2D T1-CE, 3D T1-CE

#### 2. Notation and missing-sequence handling

For patient $i$, let $S_{i}$ denote the set of available eligible sMRI sequences:

$$S_{i}\subseteq\{\text{2D FLAIR}, \text{3D FLAIR}, \text{3D T1-w}, \text{2D T1-CE}, \text{3D T1-CE}\}.$$

Let $t\in\{\mathrm{FLAIR},T1\text{-}w,T1\text{-}\mathrm{CE}\}$ index sequence types, and let $S_{i,t}$ denote the non-empty subset of available sequences from type $t$ for patient $i$. The set of available sequence types was:

$$T_{i}=\{t :\left| S_{i,t} \right|>0\}.$$

Fusion was performed only over available sequences and available sequence types.

#### 3. Sequence-level temperature scaling

Before patient-level fusion, raw sequence-level logits were calibrated by sequence-specific temperature scaling^39^ fitted exclusively on the internal validation set. For each eligible sequence subtype $q$, the temperature parameter $\tau_{q}$ was initialised at 1.5 and fitted by minimising validation-set negative log-likelihood:

$$\hat{\tau}_{q}=\arg\min_{\tau>0}\sum_{(i,s)\in\mathcal{V}_{q}} \mathrm{BCE}\left( \sigma\left( \frac{\mathcal{z}_{i,s}}{\tau} \right),y_{i} \right),$$

where $\mathcal{V}_{q}$ denotes validation samples of sequence type $q$ and $\sigma\left( \cdot\right)$ is the sigmoid function. Optimisation used L-BFGS with learning rate 0.03 and a maximum of 100 iterations.

The fitted temperatures were $\tau_{\text{3D}\text{ }\text{FLAIR}}=1.73$, $\tau_{\text{2D}\text{ }\text{FLAIR}}=1.82$, $\tau_{\text{3D}\text{ }\text{T1-CE}}=1.81$, $\tau_{\text{3D}\text{ }\text{T1-w}}=1.29$, and $\tau_{\text{2D}\text{ }\text{T1-CE}}=1.87$. In the fusion equations below, $\tau_{s}$ denotes the fitted temperature associated with sequence $s$.

#### 4. Hierarchical logit fusion

Patient-level predictions were generated using the same hierarchical logit-fusion notation as in the main Methods. Calibrated logits were first averaged within each available sequence type:

$$\mathcal{z}_{i,t}=\frac{1}{\left| S_{i,t} \right|}\sum_{s\in S_{i,t}} \frac{\mathcal{z}_{i,s}}{\tau_{s}} .$$

The resulting sequence-type logits were then averaged across available sequence types to obtain the patient-level logit:

$\mathcal{z}_{i}=\frac{1}{\left| T_{i} \right|}\sum_{t\in T_{i}} \mathcal{z}_{i,t}$ .

The final patient-level MS probability was:

$$p_{i}=\sigma\left( \mathcal{z}_{i} \right)=\frac{1}{1+\exp\left( -\mathcal{z}_{i} \right)}.$$

This two-level averaging prevented sequence types with multiple acquisitions from dominating the prediction while allowing inference when only a subset of eligible sMRI sequences was available.

#### 5. Intended end users and required expertise

DeepMS was developed for retrospective research evaluation. Intended users are imaging researchers and clinicians in research settings, with standard expertise in brain MRI interpretation for sequence/quality verification. Prospective validation and clinician-led workflow integration would be required before clinical deployment.

### S9. Interpretation analysis

#### 1. Heatmap normalisation and visualisation

For each available input sequence$s$from patient $i$, DeepMS produced a voxel-wise prediction heatmap $M_{i,s}^{\text{pred}}$ from the ABMIL interpretation branch defined in Supplementary Methods S6:

$$M_{i,s}^{\text{pred}}\left( v \right)=w^{\top}R_{i,s}\left( v \right).$$

To enable comparison across patients and sequences, the heatmap was normalised by its maximum absolute value inside the brain mask:

$$\tilde{M}_{i,s}^{\text{pred}}\left( v \right)=\frac{M_{i,s}^{\text{pred}}\left( v \right)}{\max_{u\in\Omega_{i,s}^{\text{brain}}} \left| M_{i,s}^{\text{pred}}\left( u \right) \right|+\epsilon},$$

where $\Omega_{i,s}^{\text{brain}}$ denotes the brain mask in the preprocessed input space and $\epsilon={10}^{-6}$is a numerical-stability constant. This normalisation maps heatmap values to approximately $[-1, 1].$ For qualitative visualisation, $\tilde{M}_{i,s}^{\text{pred}}$ was overlaid on the corresponding sMRI volume using the DivergingBlueRed colour map.

For activation-based analyses, we derived a binary positive activation map from the normalised prediction heatmap using the fixed threshold $\tau_{H}=0.50$:

$$A_{i,s}^{+}\left( v \right)1=\left[ \tilde{M}_{i,s}^{\text{pred}}\left( v \right)>\tau_{H} \right].$$

Group-level activation-probability maps were generated by averaging binary activation maps voxel-wise across MS cases with available lesion masks:

$$P_{s}^{+}\left( v \right)=\frac{1}{N_{s}}\sum_{i=1}^{N_{s}} A_{i,s}^{+}\left( v \right).$$

where $N_{s}$ is the number of MS cases with sequence $s$ and available lesion masks. $P_{s}^{+}\left( v \right)$ therefore represents the proportion of cases with suprathreshold positive activation at voxel $v$ for sequence $s$.

#### 2. Lesion, perilesion, and NAWM activation metrics

To assess whether lesion and perilesional signals were attenuated in the lesion-masking experiments, we quantified the residual extent of strong positive heatmap activation within conservative lesion-centric regions. This analysis was restricted to external MS cases with lesion masks.

Let $Mask_{i}^{\text{lesion}}$ denote the binary lesion mask for patient $i$. Because lesion-masking preprocessing removed voxels within a dilation radius of $d=3$ voxels, we used a stricter dilation radius of $d=4$ voxels to define the lesion-and-perilesion region of interest:

$$Mask_{i}^{\text{LP}}=\mathrm{Dilate} \left( Mask_{i}^{\text{lesion}},d=4 \right).$$

Lesion-and-perilesion activation volume was computed as a suprathreshold voxel count:

$$A_{i,s}^{\text{LP}}=\sum_{v\in\Omega_{i,s}^{\text{brain}}} A_{i,s}^{+}\left( v \right) Mask_{i}^{\text{LP}}\left( v \right).$$

For analyses of NAWM activation, the NAWM mask was defined as the brain mask excluding the lesion-and-perilesion region:

$$Mask_{i,s}^{\text{NAWM}}=\Omega_{i,s}^{\text{brain}}\setminus Mask_{i}^{\text{LP}},$$

and the corresponding NAWM activation volume was:

$$A_{i,s}^{\text{NAWM}}=\sum_{v\in\Omega_{i,s}^{\text{brain}}} A_{i,s}^{+}\left( v \right) Mask_{i,s}^{\text{NAWM}}\left( v \right).$$

All activation volumes were reported as voxel counts.

**3. Activation-lesion overlap and mean Dice**

To quantify the spatial overlap between model activation and focal lesions, we computed case-level Dice coefficients in the public external MS cases with available expert lesion masks. The binary activation maps used for this analysis were the same $A_{i,s}^{+}$.

For two binary voxel sets $B$ and $C$ within the preprocessed brain mask, Dice overlap was defined as:

$$\mathrm{Dice}_{i,s} \left( B,C \right)=\frac{2\sum_{v\in\Omega_{i,s}^{\text{brain}}} B\left( v \right)C\left( v \right)}{\sum_{v\in\Omega_{i,s}^{\text{brain}}} B\left( v \right)+\sum_{v\in\Omega_{i,s}^{\text{brain}}} C\left( v \right)+\epsilon_{D}},$$

where $\epsilon_{D}={10}^{-6}$ was added for numerical stability. We then computed case-level lesion and lesion-and-perilesion overlap as:

$D_{i,s}^{\text{lesion}}=\mathrm{Dice}_{i,s} \left( A_{i,s}^{+},Mask_{i}^{\text{lesion}} \right),$ $D_{i,s}^{\text{LP}}=\mathrm{Dice}_{i,s} \left( A_{i,s}^{+},Mask_{i}^{\text{LP}} \right).$

$Mask_{i}^{\text{LP}}$ denotes the dilated lesion-and-perilesion mask defined above. Case-level Dice values were retained for paired model comparisons. Mean Dice was obtained by averaging case-level Dice values across external MS cases with available lesion masks:

$mDice_{s}^{\text{lesion}}=\frac{1}{N_{s}}\sum_{i=1}^{N_{s}} D_{i,s}^{\text{lesion}}$, $mDice_{s}^{\text{LP}}=\frac{1}{N_{s}}\sum_{i=1}^{N_{s}} D_{i,s}^{\text{LP}}.$

**4. Effect of lesion masking**

We quantified lesion-masking effects by computing the mean number of suprathreshold heatmap voxels within the lesion-and-perilesion ROI, $A_{i,s}^{\text{LP}}$, across selected true-positive MS cases for FLAIR before and after lesion masking (Supplementary Fig. 3). Mean lesion-and-perilesion activation decreased markedly after lesion masking across all training settings: FLAIR-only, 8,343.60 to 23.79 (-99.7%); FLAIR+T1, 14,775.07 to 91.00 (-99.4%); and training with both dMRI and sMRI, 6,060.79 to 75.74 (-98.8%).

**5. Output storage and auditability**

All model outputs, including scan-level predictions and derived interpretation maps, were stored on secure institutional servers with access controls. Outputs were linked to version-controlled code, fixed preprocessing pipelines, and archived model checkpoints to support audit and reproduction. Derived outputs can be made available for evaluation upon reasonable request and appropriate approvals; individual-level imaging data for the internal and Krakow external cohorts are not publicly available because of privacy and institutional restrictions.

### S10. Calibration analysis

Patient-level probabilistic calibration was assessed for the final fused MS probabilities using calibration curves (Supplementary Fig. 4) and standard summary metrics, including the Brier score, calibration intercept, and calibration slope (Supplementary Table 7). Sequence-level temperature scaling was used only to harmonise logits across sequence types before fusion; no additional cohort-level recalibration was performed. Brier scores were low across the internal test, Krakow external, and public external cohorts (0.058, 0.057, and 0.036, respectively), whereas calibration intercepts and slopes varied across cohorts (Supplementary Table 7), indicating that patient-level probabilities may require cohort-specific interpretation when transferred across settings.

### S11. Subgroup analyses

We performed prespecified subgroup analyses by age and sex in the internal test, Krakow external, and public external cohorts. Performance was summarised using ROC-AUC with 95% CIs, computed within each stratum using the same inference and fusion pipeline as the main analysis (Extended Data Fig. 2).

#### 1. Age-stratified performance

Age subgroup analyses used prespecified strata of <40 years, 40–60 years, and >60 years. Performance was summarised using ROC curves and ROC-AUC with 95% CIs within each cohort (Extended Data Fig. 2). In the internal test cohort, ROC-AUCs were 0.967 (0.925–0.986) for <40 years, 0.972 (0.945–0.994) for 40–60 years, and 0.944 (0.828–0.999) for >60 years (overall 0.968 (0.947–0.987)). In the Krakow external cohort, ROC-AUCs were 0.907 (0.834–0.969), 0.966 (0.904–1.000), and 0.975 (0.903–1.000) across the same age strata (overall 0.940 (0.898–0.974)). In the public external cohort, ROC-AUCs were 0.963 (0.948–0.976), 0.981 (0.967–0.991), and 0.966 (0.923–0.998) (overall 0.974 (0.966–0.982)). Across cohorts, age-stratified ROC curves showed similar discrimination with overlapping CIs, noting wider uncertainty in smaller strata.

#### 2. Sex-stratified performance

Sex subgroup analyses were performed using female and male strata (cases with missing sex were excluded from this analysis). Performance was summarised using ROC curves and ROC-AUC with 95% CIs within each cohort (Extended Data Fig. 2). In the internal test cohort, ROC-AUCs were 0.967 (0.941–0.986) for females and 0.967 (0.914–0.999) for males (overall 0.968 (0.947–0.987)). In the Krakow external cohort, ROC-AUCs were 0.942 (0.892–0.980) for females and 0.936 (0.851–0.990) for males (overall 0.940 (0.898–0.974)). In the public external cohort, ROC-AUCs were 0.975 (0.959–0.988) for females and 0.982 (0.967–0.995) for males (overall 0.974 (0.966–0.982)). Across cohorts, sex-stratified ROC curves showed similar discrimination with overlapping CIs, noting wider uncertainty in smaller strata.

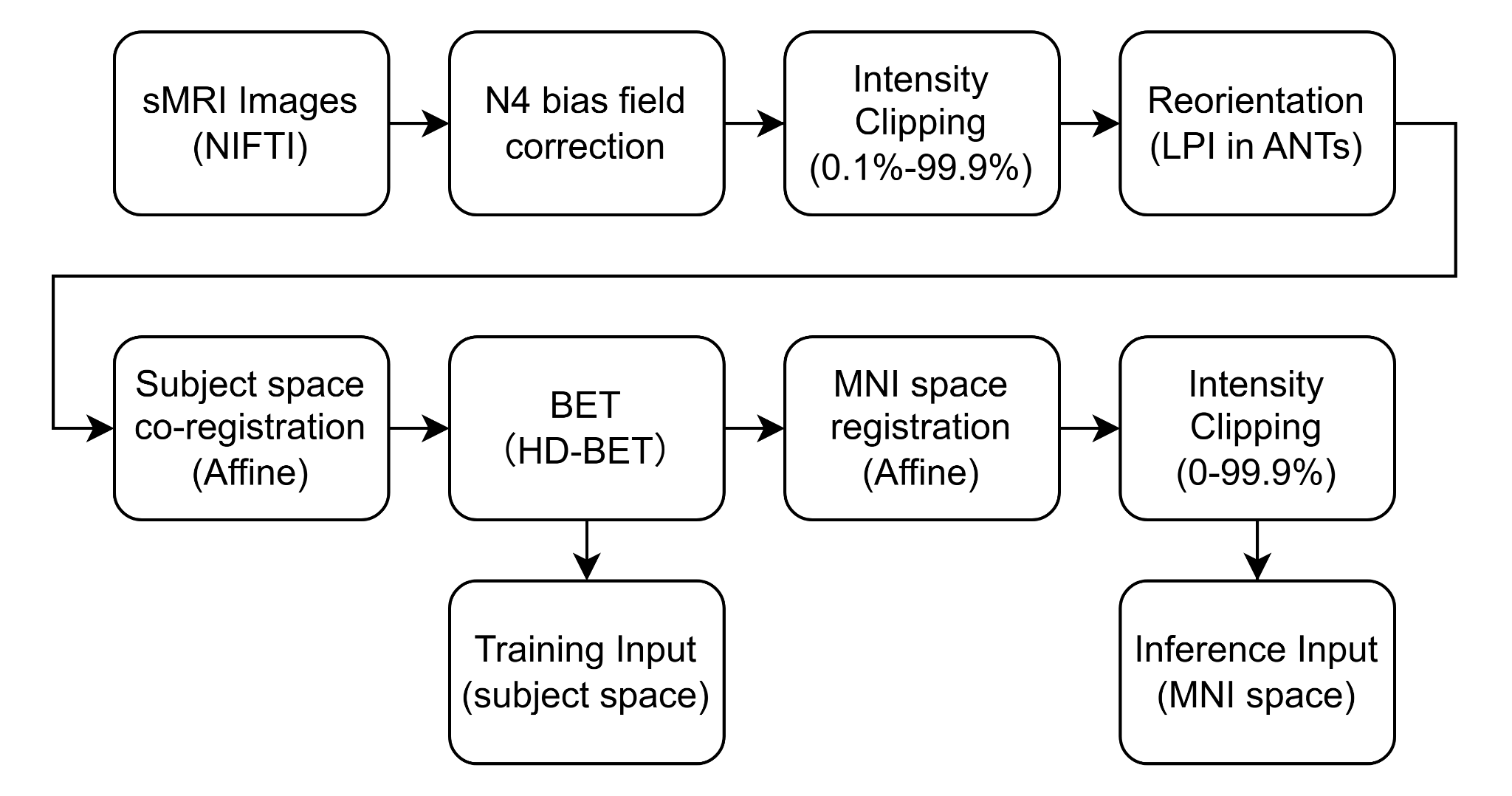

### Supplementary Fig. 1 | Structural MRI preprocessing pipeline.

Raw sMRI volumes (NIfTI format) undergo N4 bias-field correction, robust intensity clipping (0.1–99.9th percentile), and reorientation to LPI using ANTs. Images are then affinely co-registered in subject space and skull-stripped using HD-BET. These native subject-space images are used as training inputs to improve generalisation. For inference, skull-stripped images are further affinely registered to MNI space, followed by a second intensity clipping (0–99.9th percentile) to generate MNI-normalised inference inputs.

ANTs=Advanced Normalization Tools. HD-BET=high-definition brain extraction. LPI=left–posterior–inferior. MNI=Montreal Neurological Institute. N4=N4 bias-field correction. NIfTI=Neuroimaging Informatics Technology Initiative. sMRI=structural magnetic resonance imaging.

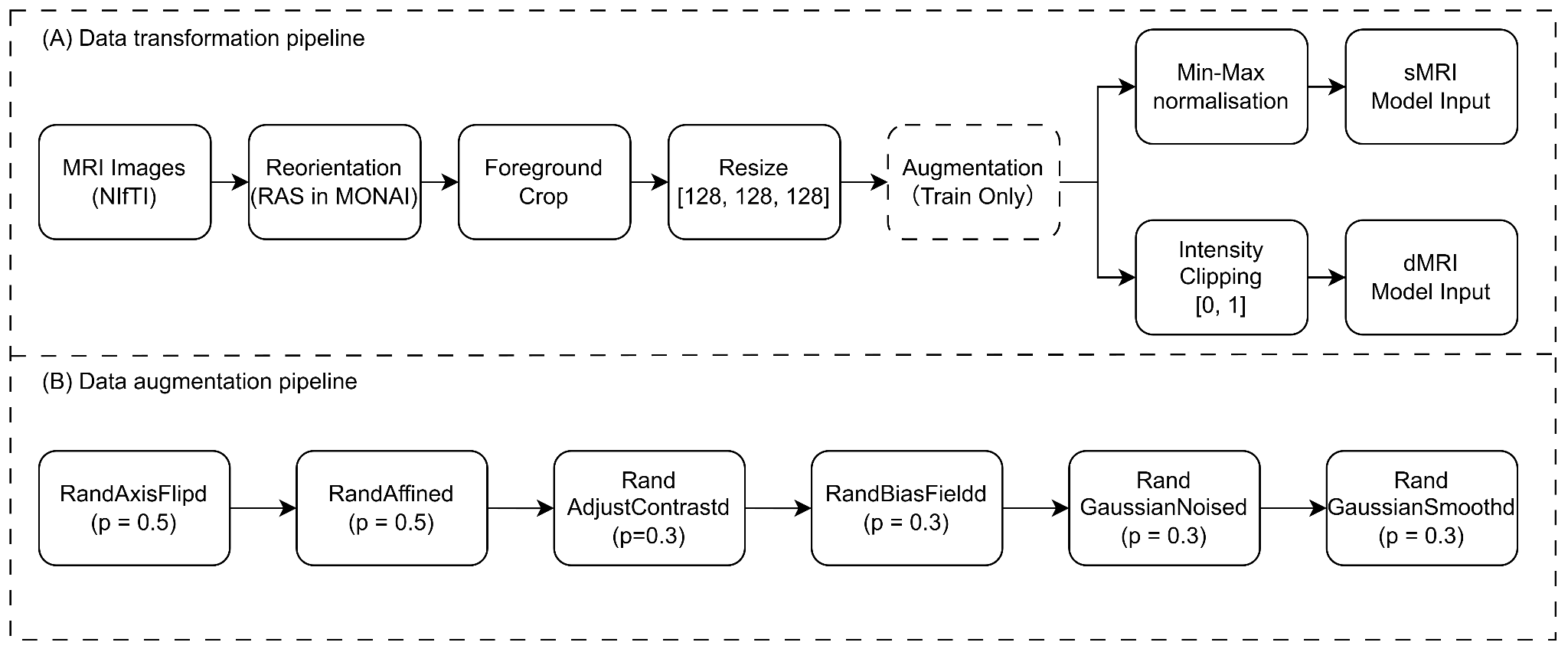

### Supplementary Fig. 2 | Data preprocessing and augmentation workflows.

**(A)** Data transformation pipeline. Raw NIfTI volumes were reoriented to the canonical right–anterior–superior (RAS) coordinate system using MONAI, foreground-cropped, and resized to 128×128×128 voxels. Geometric transformations were applied jointly to images and corresponding masks (WM and lesion masks) to maintain spatial alignment. Intensity normalisation was modality-specific: sMRI volumes underwent min–max normalisation to [0, 1], while dMRI-derived maps were intensity-clipped to the [0, 1] range. **(B)** Data augmentation pipeline is applied during training. The workflow included geometric augmentations (random axis flipping and affine transformations; p=0.5) and intensity augmentations (random contrast adjustment, random bias field, Gaussian noise, and Gaussian smoothing; p=0.3) to simulate acquisition variability.

dMRI=diffusion MRI. MONAI=Medical Open Network for AI. NIfTI=Neuroimaging Informatics Technology Initiative. p=probability of augmentation application. RAS=right–anterior–superior. sMRI=structural MRI. WM=white matter.

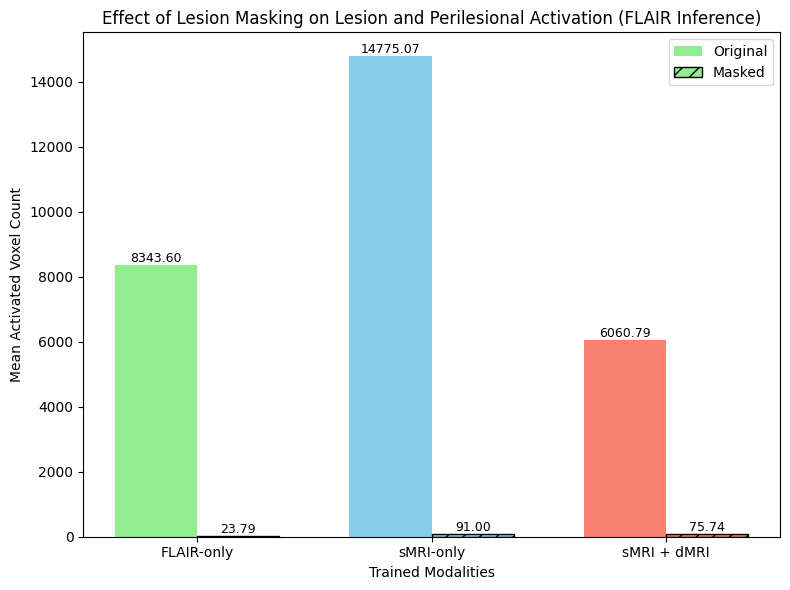

### Supplementary Fig. 3 | Effect of lesion masking on model activation in lesion and perilesional regions.

The bar chart compares the mean activated voxel counts for models trained on different modality combinations (FLAIR-only, sMRI-only, and sMRI+dMRI) in patients with multiple sclerosis. Light-coloured bars represent activation with original inputs, while hatched bars represent activation after masking the lesion and perilesional areas. The substantial reduction in voxel counts across all modalities following masking indicates the successful removal of lesion-associated diagnostic information. dMRI=diffusion MRI.

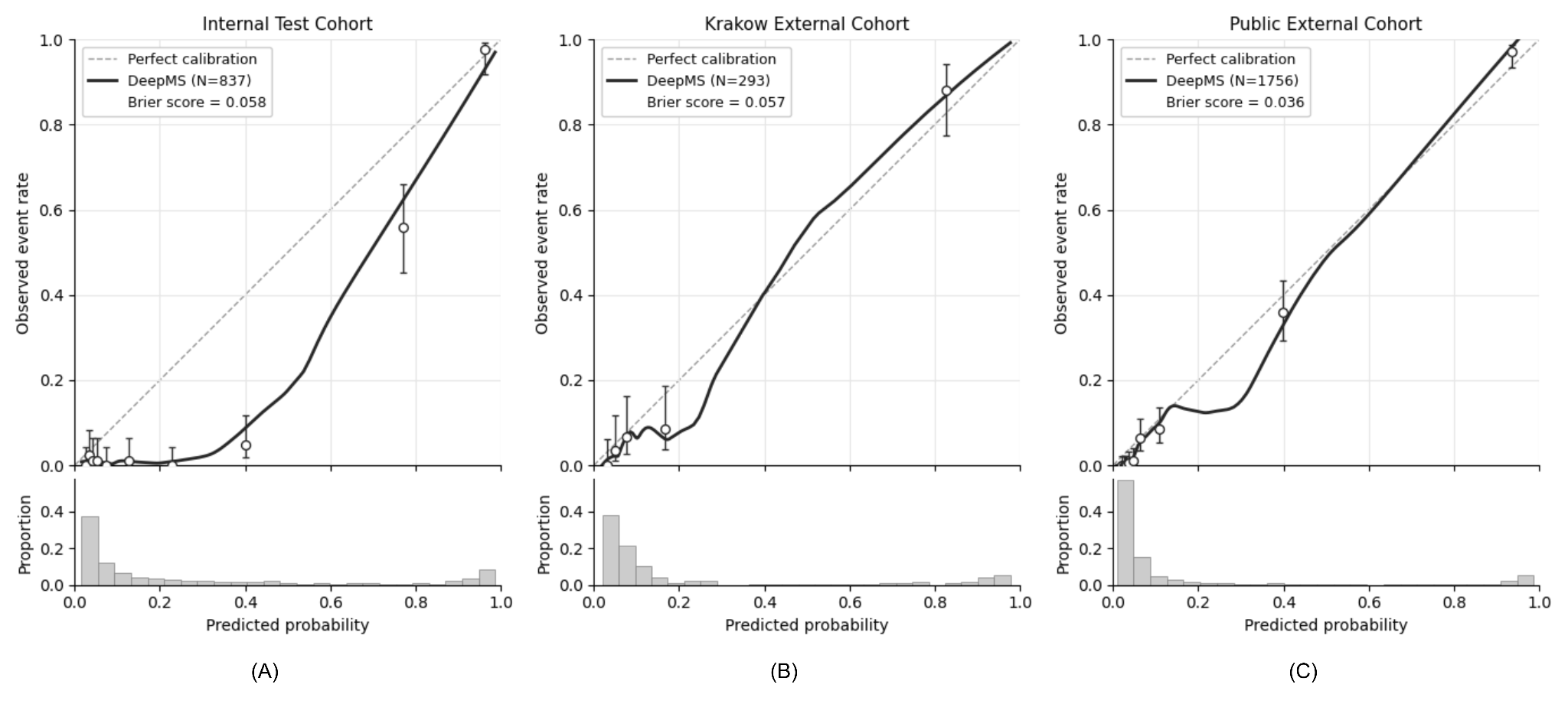

### Supplementary Fig. 4 | Calibration curves of the DeepMS model across internal and external cohorts.

The calibration plots illustrate the agreement between the predicted probability of multiple sclerosis (x-axis) and the observed event rate (y-axis) for the (A) internal test cohort (N=837), (B) Krakow external cohort (N=293), and (C) public external cohort (N=1,756).

### Supplementary Table 1 | Model performance in each public external dataset

| **Dataset** | **N (MS / Non-MS)** | **Sensitivity (95% CI)** | **Specificity (95% CI)** |
| --- | --- | --- | --- |
| **MSSEG-2016** | 53 (53/0) | 0.679 (0.553–0.807) | NA |
| **MSSEG-2** | 40 (40/0) | 0.825 (0.697–0.936) | NA |
| **OpenMS-Long** | 15 (15/0) | 0.867 (0.667–1.000) | NA |
| **OpenMS-Cross** | 27 (27/0) | 0.815 (0.656–0.952) | NA |
| **MS-ISBI** | 19 (19/0) | 0.842 (0.650-1.000) | NA |
| **PediMS** | 9 (9/0) | 0.889 (0.625–1.000) | NA |
| **QSM** | 150 (50/100) | 0.870 (0.802–0.933) | 1.000 (1.000–1.000) |
| **WMH** | 170 (0/170) | NA | 0.959 (0.928-0.988) |
| **MRBrainS18** | 30 (0/30) | NA | 1.000 (1.000-1.000) |
| **ISLES-2022** | 250 (0/250) | NA | 0.984 (0.968-0.996) |
| **UCSF-PDGM** | 495 (0/495) | NA | 1.000 (1.000-1.000) |
| **MetsToBrain** | 200 (0/200) | NA | 1.000 (1.000-1.000) |
| **MPI-Leipzig** | 117 (0/117) | NA | 0.974 (0.943-1.000) |
| **OpenNeuro epilepsy** | 170 (0/170) | NA | 0.982 (0.962-1.000) |
| **PediDemi** | 11 (0/11) | NA | 1.000 (1.000-1.000) |

### Supplementary Table 2 | Sensitivity analysis of DeepMS versus biomarkers in early MS (≤3 years)

|  | **Detected / Total** | **Sensitivity (95% CI)** | **p value** |
| --- | --- | --- | --- |
| **DeepMS** | 26/27 | 0.963 (0.879–1.000) | Reference |
| **DIS** | 24/27 | 0.889 (0.758–1.000) | 0.625 |
| **DIT** | 9/27 | 0.333 (0.166–0.522) | <0.001† |
| **CVS** | 14/27 | 0.519 (0.318–0.720) | <0.001† |
| **PRL** | 3/27 | 0.111 (0.000–0.250) | <0.001† |
| **Composite Biomarker** | 17/27 | 0.630 (0.452–0.826) | 0.008† |
| **DeepMS+Lesion** | 25/27 | 0.926 (0.807–1.000) | 1.000 |

† P < 0.05 by exact McNemar’s test compared with DeepMS (threshold = 0.50)

### Supplementary Table 3 | Structural MRI acquisition parameters in internal cohort

| **Sequence** | **TR (ms)** | **TE (ms)** | **TI (ms)** | **Flip angle** | **Voxel size (mm)** |
| --- | --- | --- | --- | --- | --- |
| **3D T1-w (± contrast)** | 2100 | 2.7 | 900 | 8° | 1.0 × 1.0 × 1.0 |
| **2D T1-w  (± contrast)** | 250 | 2.5 | — | 70° | [0.5–1.0] × [0.5–1.0] × 3.0 |
| **3D FLAIR** | 6000 | 325 | 2100 | 120° | 1.0 × 1.0 × 1.0 |
| **2D FLAIR** | 9000 | 81 | 2500 | 150° | [0.5–1.0] × [0.5–1.0] × 3.0 |
| **SWI** | 28 | 20 | — | 15° | [0.8–1.0] × [0.8–1.0] × 3.0 |

All scans were acquired on 3.0 T Siemens Magnetom Prisma and Skyra scanners.

### Supplementary Table 4 | Diffusion MRI acquisition parameters in internal cohort

| **Parameter** | **Value** |
| --- | --- |
| **Scanner** | 3.0 T Siemens Prisma / Skyra |
| **Sequence type** | Single-shot spin-echo EPI |
| **Protocol  (# direction × b-values)** | 5 × b = 0; 4 × b = 250; 20 × b = 1000; 60 × b = 2000 s/mm² |
| **TR** | 3.2–4.0 s (Prisma); 3.5–4.3 s (Skyra) |
| **TE** | 70–96 ms (Prisma); 95–100 ms (Skyra) |
| **Voxel size** | 1.7 × 1.7 × 3.0 mm³ (50 slices) |
| **Partial Fourier** | 6/8 |
| **Scanning time** | ~7 min |

All scans were acquired on 3.0 T Siemens Magnetom Prisma and Skyra scanners.

### Supplementary Table 5 | Structural MRI acquisition parameters in Krakow external cohort

| **Sequence** | **TR (ms)** | **TE (ms)** | **TI (ms)** | **Flip angle** | **Voxel size (mm)** |
| --- | --- | --- | --- | --- | --- |
| **3D T1-w (± contrast)** | 8.5 | 3.6 | 450 | 12° | [0.4-0.6] × [0.4-0.6] × [1.0-1.6] |
| **3D FLAIR** | 6200 | 90 | 1760 | 90° | [0.4-0.6] × [0.4-0.6] × [1.0-1.6] |

All scans were acquired on 1.5 T GE SIGNA Artist scanners.

### Supplementary Table 6 | Summary of implementation details and hyperparameters.

| Parameter / Setting | Value / Details |
| --- | --- |
| **Data & augmentation** |  |
| Input modalities | sMRI (FLAIR, T1-w, T1CE, $b=0$) + dMRI maps (DTI/DKI/SMI) |
| Input size | $128 \times128 \times128$ |
| Augmentation | Random Axis Flip (p=0.5), Random Affine (rot ±5°, trans ≤5 voxels, p=0.5),  Random Contrast Adjustment (gamma range 0.7-1.3; p=0.3), Bias-field Artefacts (p=0.3), Additive Gaussian Noise (p=0.3), and Gaussian Smoothing (p=0.3). |
| Sampling strategy | Stratified oversampling (label × input type) |
| **Model architecture** |  |
| Feature extractor | Swin-UNETR (Init Weights: “VoComni-B”) |
| Prediction head | ABMIL (2 heads) |
| Classifier output | Single logit (Sequence level binary MS classification) |
| **Training & optimisation** |  |
| Loss functions | $\mathcal{L}_{\text{total}}=\underset{\mathcal{L}_{\text{cls}}}{\underbrace{\text{BCEWithLogits}\left( \mathcal{z}_{i,s},y_{i} \right)}}+\underset{\mathcal{L}_{\text{non-brain}}}{\lambda_{\text{nb}}\underbrace{\parallel M_{i,s}^{\text{reg}}\odot Mask_{i,s}^{\text{non-brain}}\parallel_{1}}}+\underset{\mathcal{L}_{\text{neg}}}{\lambda_{\text{neg}}\underbrace{1\left[ y_{i}=0 \right]\parallel\text{ReLU}\left( M_{i,s}^{\text{reg}} \right)\parallel_{1}}}$ |
| Loss weights | $\lambda_{\text{nb}}={10}^{-6};$  $\lambda_{\text{neg}}=0$ for epochs 1-4, linearly increased from ${10}^{-8}$ to ${10}^{-7}$ during epochs 5-15, and set to 0 thereafter. |
| Optimiser | AdamW (Init LR: ${10}^{-4}$, Cosine decay) |
| Batch size | 16 |
| Training duration | Max 40 epochs (Early stopping patience = 5) |
| Precision | AMP (FP16) |
| Random seed | 42 |

### Supplementary Table 7 | Calibration analysis of DeepMS in different cohorts

|  | **MS / Total** | **Brier score** | **Calibration intercept** | **Calibration slope** |
| --- | --- | --- | --- | --- |
| **Internal Test** | 138 / 837 | 0.058  (0.050 – 0.068) | -1.435  (-1.652 – -1.242) | 1.479  (1.260 – 1.822) |
| **Krakow External** | 63 / 293 | 0.057  (0.037 – 0.077) | -0.201 (-0.579 – 0.130) | 1.221  (0.956 – 1.701) |
| **Public External** | 263 / 1756 | 0.036  (0.030 – 0.042) | -0.363 (-0.535 – -0.203) | 1.365  (1.238 – 1.527) |

### Supplementary Table 8 | Summary of prior MRI-based AI studies for multiple sclerosis diagnosis

| Study (Year) | Study Cohort | Modality | Model type | Performance |
| --- | --- | --- | --- | --- |
| Eitel et al.  (2019)^40^ | Single centre MS: 76 patients HC: 71 patients  Train+Val: Test = 8.5:1.5 | 3D FLAIR | CNN | Internal Test  - ACC: 87.04%  - ROC-AUC: 96.08%  - Sensitivity: 93.08%  - Specificity: 81.00% |
| Wang et al.  (2020)^41^ | Single centre MS: 47 patients  NMOSD: 41 patients | 3D FLAIR | ResNet18 | Mean Internal Val  (5-Fold CV)  - ACC: 75.0%  - Sensitivity: 70.7%  - Specificity: 75.9% |
| Lopatina et al.  (2020)^42^ | Single centre  MS: 66 patients  HC: 66 patients Train+Val: Test = 1:1 | SWI | CNN | Internal Test  - ACC: 92% |
| Kim et al.  (2020)^43^ | Single centre  MS: 213 NMOSD: 125   Train: Val: Test = 45:15:40 | 2D FLAIR + Clinical Info | ResNeXt | Internal Test  - ACC: 71.1%  - ROC-AUC: 82%  - Sensitivity: 61.6%  - Specificity: 87.8% |
| Hagiwara et al.  (2021)^44^ | Single centre  MS: 35 patients  NMOSD: 18 patients | R1, R2, and proton density maps | CNN | Leave-one-out CV  - ACC: 81.1%  - ROC-AUC: 85.9%  - Sensitivity: 80.0% |
| Ekşi et al.  (2021)^45^ | Internal Data MS: 51 patients  INTERPRET dataset Low grade tumor: 39  Train: Test = 7:3 | MRS | Feature Engineering + ANN | Internal Test  - ACC: 100%  - Sensitivity: 100%  - Specificity: 100% |
| Rocca et al.  (2021)^46^ | Single centre (Train+Val/Test)  MS: 70 (52/18)  Migraine: 56 (34/22)  NMOSD: 91 (56/35)  CNS vasculitis: 51 (36/15) | 2D/3D T1-w, 2D/3D T2-w | 3DCNN | Internal Test  - MS ACC: 98.8%  - Migraine ACC: 92.9%  - NMOSD ACC: 88.6%  - Vasculitis ACC: 92.1% |
| Huang et al.  (2022)^47^ | Single centre Train + Val MS: 53 patients NMOSD: 129 patients MOGAD: 49 patients  Test MS: 14 patients NMOSD: 33 patients MOGAD: 12 patients | 2D FLAIR, 2D T2-w | Transformer (Slice input) + multiple instance learning | Internal Test  - ACC: 81.4%  - ROC-AUC: 93.3% |

(Continue)

| Study (Year) | Study Cohort | Modality | Model type | Performance |
| --- | --- | --- | --- | --- |
| Seok et al.  (2023)^48^ | Single centre  MS: 86  NMOSD: 70 | 3D FLAIR (5 axial slices) | ResNet-18 | Mean Internal Val  (100 random resplit)  - ACC: 76.1%  - ROC-AUC: 85%  - Sensitivity: 77.3%  - Specificity: 74.8% |
| Khattap et al.  (2024)^49^ | Development cohort  (1 public dataset)  MS: 38 patients  HC: 38 patients  Test Cohort (Single centre)  MS: 262 patients  HC: 163 patients | 2D FLAIR, or 2D T2-w  (Slice Image) | Radiomics + Random Forest | Test Cohort  (Image level)  - ACC: 91.24% (T2-w)  - ACC: 92.94% (FLAIR) |
| Amin et al.  (2024)^50^ | Single centre  MS: 250 scans  Non-specific WMLs: 250 scans | 3D FLAIR, 2D T2-w, 3D T1-w | CNN | Mean Internal Test (5-fold CV)  - ACC: 78%  - Sensitivity: 86%  - Specificity: 72% |
| Xu et al.  (2024)^51^ | Single centre  MS: 112 patients cSVD: 321 patients  Train: Val: Test = 7:1:2 | 2D FLAIR | ResNet34 + Dual Attention Modules | Mean Internal Test  (5 random data resplits)  - ACC: 86.06%  - ROC-AUC: 98.78%  - Sensitivity: 96.89%  - Specificity: 89.17% |
| Huang et al.  (2024)^52^ | Single centre  MS: 48 patients NMOSD: 62 patients  1 Public dataset  MS: 21 patients | 3D FLAIR | Swin-Transformer + CNN | Mean Internal Val  (5-Fold CV) - ACC: 92.36% |
| Cortese et al. (2025)^53^ | MAGNIMS Multicentre  (Training/Testing)  MS: 150 (120/30)  MOGAD: 115 (92/23)  Independent Validation  MS: 68  MOGAD: 73 | 3D FLAIR, 3D T1-w, Clinical Info | ResNet-10 | Independent Validation  - ACC: 86%  - ROC-AUC: 90%  - Sensitivity: 84%  - Specificity: 89% |

ACC=accuracy; ANN=artificial neural network; CNN=convolutional neural network; CV=cross-validation; FLAIR=fluid-attenuated inversion recovery; HC=healthy controls; MOGAD=myelin oligodendrocyte glycoprotein antibody–associated disease; MRS=magnetic resonance spectroscopy; MS=multiple sclerosis; NMOSD=neuromyelitis optica spectrum disorder; ROC-AUC=area under the receiver operating characteristic curve; SWI=susceptibility-weighted imaging; T1-w=T1-weighted; T2-w=T2-weighted; WML(s)=white matter lesion(s); cSVD=cerebral small vessel disease.

### STARD 2015 Checklist

|  | **Section & Topic** | **No** | **Item** | **Reported on page #** |
| --- | --- | --- | --- | --- |
|  | **TITLE OR ABSTRACT** |  |  |  |
|  |  | **1** | Identification as a study of diagnostic accuracy using at least one measure of accuracy  (such as sensitivity, specificity, predictive values, or AUC) | Main pp 1 |
|  | **ABSTRACT** |  |  |  |
|  |  | **2** | Structured summary of study design, methods, results, and conclusions  (for specific guidance, see STARD for Abstracts) | Main pp 1 |
|  | **INTRODUCTION** |  |  |  |
|  |  | **3** | Scientific and clinical background, including the intended use and clinical role of the index test | Main pp 2-4 |
|  |  | **4** | Study objectives and hypotheses | Main pp 3-4 |
|  | **METHODS** |  |  |  |
|  | *Study design* | **5** | Whether data collection was planned before the index test and reference standard  were performed (prospective study) or after (retrospective study) | Main pp 4, 14-15 |
|  | *Participants* | **6** | Eligibility criteria | Main pp 14-15 |
|  |  | **7** | On what basis potentially eligible participants were identified  (such as symptoms, results from previous tests, inclusion in registry) | Main pp 14-15 |
|  |  | **8** | Where and when potentially eligible participants were identified (setting, location and dates) | Main pp 14-15 |
|  |  | **9** | Whether participants formed a consecutive, random or convenience series | Main pp 14-15 |
|  | *Test methods* | **10a** | Index test, in sufficient detail to allow replication | Main pp 16-20  Supplementary pp 8-15 |
|  |  | **10b** | Reference standard, in sufficient detail to allow replication | Main p 21 |
|  |  | **11** | Rationale for choosing the reference standard (if alternatives exist) | Main pp 2-4, 21 |
|  |  | **12a** | Definition of and rationale for test positivity cut-offs or result categories  of the index test, distinguishing pre-specified from exploratory | Main p 22-23 |
|  |  | **12b** | Definition of and rationale for test positivity cut-offs or result categories  of the reference standard, distinguishing pre-specified from exploratory | Main p 22-23 |
|  |  | **13a** | Whether clinical information and reference standard results were available  to the performers/readers of the index test | Main pp 16-20 |
|  |  | **13b** | Whether clinical information and index test results were available  to the assessors of the reference standard | Main p 21 |
|  | *Analysis* | **14** | Methods for estimating or comparing measures of diagnostic accuracy | Main pp 22-23 |
|  |  | **15** | How indeterminate index test or reference standard results were handled | Main p 15 |
|  |  | **16** | How missing data on the index test and reference standard were handled | Main pp 15, 21 |
|  |  | **17** | Any analyses of variability in diagnostic accuracy, distinguishing pre-specified from exploratory | Main p 6, Supplementary p 18 |
|  |  | **18** | Intended sample size and how it was determined | No formal sample size calculation was performed because this retrospective study used a consecutive (all eligible) sample. |
|  | **RESULTS** |  |  |  |
|  | *Participants* | **19** | Flow of participants, using a diagram | Main pp 14-15, Extended Data Fig. 1 |
|  |  | **20** | Baseline demographic and clinical characteristics of participants | Main pp 4-5, Table 1, Extended Data Table 1-2 |
|  |  | **21a** | Distribution of severity of disease in those with the target condition | Main p 7  Supplementary p 23 |
|  |  | **21b** | Distribution of alternative diagnoses in those without the target condition | Main pp 4-5, Table 1, Extended Data Table 1-2 |
|  |  | **22** | Time interval and any clinical interventions between index test and reference standard | Main p 21, the same MRI scan was used |
|  | *Test results* | **23** | Cross tabulation of the index test results (or their distribution)  by the results of the reference standard | Main pp 5-6, Table 2; |
|  |  | **24** | Estimates of diagnostic accuracy and their precision (such as 95% confidence intervals) | Main pp 5-6, Table 2 |
|  |  | **25** | Any adverse events from performing the index test or the reference standard | No adverse events were observed or expected. |
|  | **DISCUSSION** |  |  |  |
|  |  | **26** | Study limitations, including sources of potential bias, statistical uncertainty, and generalisability | Main p 13 |
|  |  | **27** | Implications for practice, including the intended use and clinical role of the index test | Main pp 11-14 |
|  | **OTHER INFORMATION** |  |  |  |
|  |  | **28** | Registration number and name of registry | Not registered. |
|  |  | **29** | Where the full study protocol can be accessed | Main p 23 (design, code for training, validation and testing) |
|  |  | **30** | Sources of funding and other support; role of funders | Main p 24 |
